# Uncovering identifiability of epidemiological models: basic reproduction number and complementary data streams

**DOI:** 10.64898/2026.01.16.26344284

**Authors:** Binod Pant, Omar Saucedo, Gleb Pogudin

## Abstract

Mathematical models of infectious disease dynamics are routinely fitted to surveillance data to estimate epidemiological parameters and inform public health decisions. Such data are typically discrete and noisy, but before attempting estimation, it is essential to ask whether the model structure itself permits unique parameter identification at least under perfect (continuous, noise-free) observations. This mathematical property of a model with respect to observation(s), known as structural identifiability, serves as a necessary precondition for reliable inference, since a model that fails this test cannot yield unique parameter estimates even from perfect data. In this study, we systematically investigate structural identifiability in various classes of compartmental epidemic models and establish two main findings. First, we present and deploy a methodology for assessing structural identifiability of epidemiological quantities of interest and demonstrate that the basic reproduction number exhibits identifiability across diverse model structures—including models with multiple transmission pathways and host-vector dynamics—even when individual parameters are not uniquely identifiable. These findings challenge the assumption that complete model identifiability is necessary for reliable epidemiological inference and suggest reformulating the central question from “is the model identifiable?” to “are the quantities that matter for the decision-making identifiable?” Second, we prove that incorporating minimal complementary data, as little as a single time-point measurement from an additional state variable, can make otherwise nonidentifiable models globally identifiable. This result has direct implications for surveillance design: rather than putting limited resources into frequent monitoring of multiple data streams or relying on external parameter estimates that may be uncertain or context-dependent, public health systems can strategically prioritize collecting high-quality complementary measurements.

## 1 Introduction

Mathematical models of infectious disease dynamics play a central role in public health decision-making, from forecasting epidemic trajectories to evaluating intervention strategies [12, 25, 45]. These models provide insights into key epidemiological quantities such as the peak timing and the cumulative burden of infectious individuals, all of which depend on the underlying model parameters and state variables. To generate predictions during ongoing outbreaks and enable retrospective analysis of past epidemics, model parameters are often estimated by fitting the model to observed data, typically comprising measurements such as reported cases, hospitalizations, or mortality. However, a fundamental challenge confronts this modeling pipeline: the observable data often captures only a partial view of the epidemic process. Even under ideal conditions with perfect, noise-free model-generated observations, the model structure and available measurements may not contain sufficient information to uniquely determine all parameters—a limitation formally characterized by the concept of structural identifiability [2, 14, 15, 34]. Understanding whether and when parameters can be uniquely identified from perfect observations is, therefore, essential for assessing the reliability of model-based inferences and public health decision-making.

Structural identifiability is an *a priori* theoretical property that characterizes whether the model parameters can be uniquely determined under ideal conditions: noise-free observations with no model error (i.e., observation generated from the model), where an observation is continuous for all time [2, 14, 15, 34]. It represents the best-case scenario for parameter inference, asking whether the model structure itself, in combination with the chosen observations, permits unique parameter identification. A structurally nonidentifiable model cannot produce unique parameter estimates from the given observations, regardless of data quality or quantity, making identifiability analysis an essential first step before attempting parameter estimation and uncertainty quantification [14, 33, 38, 43, 46]. Structural identifiability is thus a necessary, though not sufficient, condition for practical identifiability of parameters, where data can be noisy and discrete [26, 46, 53].

Closely related to structural identifiability is the concept of state observability [1, 15, 28, 36], which addresses whether the state variables of a differential equation can be reconstructed from observations and identified parameters. In other words, state observability concerns the identifiability of state variables rather than the model parameters. From an epidemiological perspective, the observability of state trajectories—such as the time evolution of susceptible, infectious, and recovered populations—often provides more actionable insights than the parameter values themselves. For instance, while the transmission rate (a product of the number of contacts per unit of time and transmission probability per contact, both of which depend on various factors) may be difficult to measure or interpret directly, knowing the number of currently recovered individuals has immediate implications for outbreak risk and intervention targeting. Understanding when state variables are observable, even when some parameters remain nonidentifiable, is therefore crucial for practical epidemic monitoring and control.

Beyond individual parameters and state variables, epidemiological decision-making often relies on composite quantities that aggregate information from multiple model components (e.g., a combination of parameters and state variables). The basic reproduction number, defined as the average number of secondary infections produced by a typical infectious individual in a fully susceptible population, serves as a fundamental threshold in deterministic models that determines whether an outbreak will grow or decline [51, 52]. Unlike most individual parameters, the basic reproduction number can directly inform critical public health questions: Will the disease spread? What proportion of the population must be vaccinated to prevent an outbreak? How effective must interventions be to control transmission? Despite its central importance, the structural identifiability of the basic reproduction number and similar composite quantities has received limited attention in the literature [15, 16, 30].

Despite the fundamental importance of structural identifiability and state observability, they remain understudied in epidemiological modeling. While some researchers conduct structural identifiability analysis before parameter estimation [30, 33, 43, 50], state observability is rarely examined in current epidemiological practice [36]. A critical gap exists in recognizing that structurally nonidentifiable models can still produce identifiable composite quantities or observable state variables—a straightforward yet often overlooked insight. Key questions therefore, remain: which epidemiologically relevant quantities remain identifiable or observable even when complete model identifiability fails? What structural features commonly cause non-identifiability across epidemiological model classes? What strategies achieve identifiability when models are nonidentifiable?

When epidemiological models are structurally nonidentifiable, one common strategy for achieving identifiability is to strategically fix parameters based on external data sources (such as laboratory measurements, clinical studies, or demographic surveys) such that correlations between parameters break down [49]. Alternatively, incorporating time series from a complementary data stream that provides additional information can resolve parameter unidentifiability. However, there is very little research on the minimal amount of data required from complementary streams to achieve structural identifiability of the entire system. Recently, in a computational epidemiology study, Pant et al. [43] demonstrated that fitting an SIR model to model-generated detected incidence data (where only a fraction of true incidence is detected) yields the expected non-identifiability, but incorporating even a single noise-free “ideal” seroprevalence data point reduced parameter uncertainty by orders of magnitude. Under ideal conditions–including perfect test sensitivity and specificity, representative sampling, and persistent antibody detection—such a seroprevalence measurement is equivalent to a noise-free observation of the *S*(*t*) curve [43]. Moreover, the parameter values they obtained in the noise-free setting converged to unique values, demonstrating practical identifiability and suggesting structural identifiability of the SIR model under the observation of detected incidence combined with one data point in *S*(*t*) curve. Similarly, Bergström et al. [4] recently proved that an SIR model with under-reporting and prior immunity becomes identifiable when supplemented with sample survey data of prior immunity or prevalence during the outbreak. These findings have profound implications: they suggest that even a single noise-free data point from a complementary data stream can resolve structural identifiability issues. However, this phenomenon—that minimal complementary observations can achieve global identifiability—has not, to the best of the authors’ knowledge, been systematically investigated across diverse epidemiological model classes. Understanding when and why single-time-point observations resolve structural non-identifiability has important implications for surveillance design. Rather than putting limited resources in continuously monitoring multiple data streams or relying on external parameter estimates that may be uncertain or context-dependent, public health systems can strategically prioritize collecting high-quality complementary measurements at key time points.

To summarize, this study systematically investigates structural identifiability across major epidemiological model classes with two primary objectives. First, we present and deploy a methodology for assessing the identifiability of composite epidemiological quantities, specifically the basic reproduction number. Moreover, we demonstrate that structurally nonidentifiable models can still yield identifiable epidemiologically relevant quantities, particularly the critical threshold of the basic reproduction number, challenging the assumption that complete model identifiability is necessary for reliable epidemiological inference. Second, we prove that strategic incorporation of complementary observations, often requiring as little as a single time-point measurement, can achieve global identifiability, providing a methodological theoretical foundation for minimal complementary data requirements in surveillance design.

Throughout the manuscript, we also attempt to catalog the structural mechanisms that commonly cause non-identifiability across diverse model classes and identify practical strategies for restoring identifiability. Although systematic identifiability analysis across infinite series of related models has been established for linear dynamical systems (e.g.[7, 8, 24, 37]), such investigations are rare for classes of nonlinear epidemiological models, where structural complexity can produce non-obvious patterns. While we were able to establish global identifiability for an SIR model with gamma-distributed latent period, we show that general patterns for behavioral response models with saturating incidence remain more elusive, highlighting the difficulty of deriving general identifiability results for nonlinear models.

## 2 Methods and Definitions

### 2.1 Identifiability and Observability: Definitions and Basic Properties

There exist a number of approaches to defining identifiability and observability, see [1] and [28, Section 2]. Since we deal with autonomous ODEs and our models do not have external inputs, this allows us to simplify the presentation. Below we give the definitions used throughout the paper.

Consider a dynamic model described by a system of ordinary differential equations **x**^*′*^(*t*) = **f** (**x**(*t*), ***θ***), where **x**(*t*) ∈ ℝ^*n*^ are state variables, ***θ*** ∈ ℝ^*p*^ is the parameter vector, and **y**(*t*) = **h**(**x**(*t*), ***θ***) where **y**(*t*) ∈ ℝ^*m*^ is the observation function. For fixed parameter values ***θ***^⋆^ ∈ ℝ^*p*^ and initial conditions 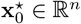, we will denote the observation of the unique analytic solution in the neighborhood of *t* = 0 by 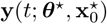.

Intuitively, identifiability means that the value of a parameter, or a state, or a function of the states and parameters, can be uniquely reconstructed from the observed data, given a “generic” observation trajectory. This can be formalized as follows.

#### Definition 1

(Global Identifiability). A function *h*(**x**(*t*), ***θ***) is called *globally identifiable* if there exists an open dense subset *U* ⊂ ℝ^*n*+*p*^ such that, for any 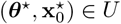, the following holds

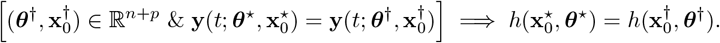

#### Example 1

Consider a basic scalar model of exponential growth *x*^*′*^(*t*) = *θ*_1_*x*(*t*) with the observation being *y*(*t*) = *θ*_2_*x*(*t*). This differential equation has a solution 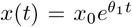, from which we get 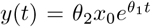. From this formula, we can deduce the following identifiability results.

- Neither *x*_0_ nor *θ*_2_ is identifiable. Indeed, for any triple 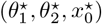 with nonzero 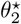 and 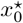, we can take a triple 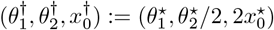 which yields the same *y*(*t*) but has 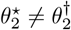 and 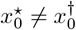.
- Parameter *θ*_1_ is globally identifiable. In order to show this, we take 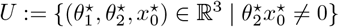. We take 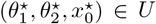 and 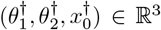. Then the equality of functions 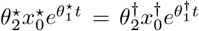 together with 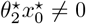 implies that 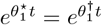, so 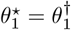. Note that this argument would not work if we were not allowed to restrict to an open dense set *U* because of special zero trajectories of the model More precisely, when *x*_0_ = 0, we have *y*(*t*) ≡ 0 regardless of the value of *θ*_1_, making *θ*_1_ nonidentifiable from such trajectories. Considering the open dense subset *U* allows us to focus on generic behavior instead of such special solutions.

#### Remark 1

(Identifiability vs. Observability). Often, in the literature, term “identifiability” is used for parameters and functions of them, while “observability” is employed for the states. We will not make such a distinction since we are sometimes interested in the identifiability of quantities involving both states and parameters. Thus, we will use the term “identifiability” for parameters and states.

#### Definition 2

(Local Identifiability). A function *h*(**x**(*t*), ***θ***) is called *locally identifiable* if there exists an open dense subset *U* ⊂ ℝ^*n*+*p*^ with the following property. For any 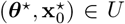, there is a neighborhood *V* of 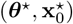 such that

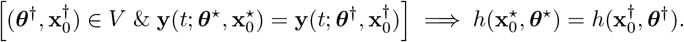

#### Example 2

We continue with the model *x*^*′*^(*t*) = *θ*_1_*x*(*t*) from Example 1 but take the observation function to be *y*(*t*) = *x*(*t*)^2^. In this case, the solution is 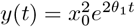, and *x*_0_ is still not identifiable. Indeed, for any pair 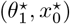 with nonzero 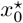, we can take a pair 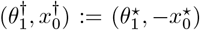 which yields the same *y*(*t*) but has 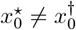.

On the other hand *x*_0_ is locally identifiable. This can be shown by taking 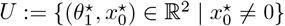 and, for every 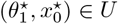 taking *V* such that 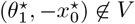. Then one can verify that no other pair in *V* yields the same *y*(*t*).

#### Remark 2

Global identifiability implies local identifiability, but the converse does not hold. In the case when the function is not even locally identifiable, we will call it *nonidentifiable*.

#### Definition 3

(Model Identifiability). A model is locally identifiable if all of its states and parameters are locally identifiable, and globally identifiable if all of its states and parameters are globally identifiable.

#### Remark 3

(Arithmetic of identifiable functions). Sums, differences, products, and quotients of globally identifiable functions of parameters are globally identifiable: one can take the intersection of the corresponding open dense sets. Furthermore, sums, differences, products, and quotients of locally identifiable functions of parameters are locally identifiable. Again, it is sufficient to take the intersections of the open dense sets *U* and the neighborhoods *V*. Note that such a combination of locally identifiable functions may be globally identifiable even if its components were not. In particular, a function of parameters which are not globally identifiable may be still identifiable.

#### Remark 4

(On the ground field). One often defines identifiability working on the field of complex numbers instead of ℝ (see, e.g., [28]) because in this case there is always a single “generic” behavior while, over reals, identifiability may be different for *θ*_*i*_ *>* 0 and *θ*_*i*_ *<* 0. Furthermore, all the software tools for assessing global identifiability/observability employ algorithms valid over complex numbers.

We have chosen to work primarily over ℝ as this is a natural choice in the context of epidemiological models. Moreover, if identifiability is established over complex numbers (using any of the available software tools), then it definitely holds over reals. For showing nonidentifiability, we will always exhibit an input-preserving transformation valid over reals as well.

### 2.2 Differential Algebra Approach to Structural Identifiability

Analyzing structural identifiability of nonlinear models often requires performing complex computations. For this reason, a number of software packages have been developed for assessing structural identifiability [47]. The analysis in this paper was carried out with the help of a software package `StructuralIdentifiability.jl` [22] (version 0.5.18 used) based on the so-called differential algebra approach. We summarize the approach used by the tool in the four steps below. For further details, we refer to Appendix A where we demonstrate all these steps on a simple SIR model.

**(Step 1)** Perform algebraic manipulations to eliminate the state variables (**x**’s) and obtain relations involving only observations, external inputs (if any), and parameters. These relations are typically called *input-output equations* of the model.

**(Step 2)** Check whether the coefficients of these relations give a complete set of identifiable parameter combinations [22, Section 5.3]; this is typically done by checking linear independence of certain monomials in the input-output equations (e.g., see Appendix A). This condition is satisfied in a majority of models in the literature [22, Table 3] and, in particular, true for all the models considered in this paper.

**(Step 3)** Check if individual parameters can be expressed in terms of these identifiable combinations [22, Section 5.2] and/or produce a simplified version of the combinations [18].

**(Step 4)** Use the identifiable combinations obtained in the previous steps together with Lie derivatives of the output to assess identifiability of state variables [18].

When performed by software, some of these steps involve *randomization*, meaning that, with some probability, the returned result may be incorrect. While the user can set any positive upper bound on the probability of the error, and in practice this probability is extremely small, we cannot use the software for all four steps if we aim at proving mathematically rigorous theorems. This issue cannot be resolved by just changing the tool: all the efficient software packages that can assess global identifiability for the states (such as SIAN [27]) are randomized.

More precisely, **(Step 1)** is deterministic, so we can use the software to rigorously compute the input-output equations. To gain additional confidence, we double-check this step using DAISY (Version 2.1) [3] in the single-output case (as in this case, the io-equations are uniquely defined). Furthermore, while **(Step 2)** involves randomization, if the software returns that the coefficients give a complete set of identifiable combinations, this is guaranteed to be true. On the other hand, **(Step 3)** and **(Step 4)** are inherently randomized, so we use them only to experiment and formulate conjectures. For the rigorous proofs, we either give a self-contained mathematical argument or, in more complex cases, employ software for computations with rational functions [17] (see, e.g., Section S4 of the Supplementary Materials). We used version 0.2.3 of the package. In order to simplify our analytic proofs for the last two steps, we also use the reparametrization functionality of the software [19]. Although the reparametrization algorithm is also randomized, the resulting variable transformation can always be checked by hand and then used safely. After performing this general identifiability analysis of a model, we use its results to characterize the impact of incorporating extra data points from a complementary data stream.

Finally, we note that not all the proofs in the paper can be performed with the help of structural identifiability software: Theorem 3.2 concerns a sequence of models of arbitrarily large dimensions, thus requiring a completely theoretical proof.

### 2.3 Visualization of Identifiability Through Multi-Start Optimization

To complement theoretical identifiability analysis, we occasionally visualize identifiability through numerical parameter estimation using multi-start local optimization, which fits the model to simulated complete outbreak data (spanning the full epidemic trajectory) from multiple initial parameter guesses distributed throughout the parameter space [43].

Structural identifiability issues manifest as non-uniqueness in the optimization landscape: globally nonidentifiable parameters yield multiple distinct parameter sets with equivalent or nearly equivalent fits to the data, while locally identifiable parameters may converge to a small discrete set of solutions. In contrast, globally identifiable parameters consistently converge to the true values regardless of the initial guess. We implement multi-start optimization in MATLAB (R2023b) using local optimizers (such as `fmincon`) with randomized initial guesses. We retain only parameter estimates that produce visually indistinguishable fits to the synthetic noise-free data, thus excluding runs that failed to converge [43].

### 2.4 Basic Reproduction Number

The basic reproduction number, ℛ_0_, represents the average number of secondary infections produced by a single infected individual introduced into a fully susceptible population. It serves as a threshold parameter in deterministic disease models: ℛ_0_ *>* 1 indicates that a disease will spread in the population, while ℛ_0_ *<* 1 typically implies that the disease will eventually clear from the population. For deterministic compartmental models, ℛ_0_ can be calculated using the next-generation matrix approach [21, 51]; we leave the details of these derivations to the reader.

## 3 Results

The main results of this study are as follows.

i. We present a methodology for assessing structural identifiability of epidemiologically relevant quantities of interest, such as the number of infectious individuals at a given point in time and the basic reproduction number ℛ_0_. We demonstrate that models, which are themselves structurally nonidentifiable can still yield structurally identifiable quantities of interest that are epidemiologically useful for decision-making. In particular, we show through various examples that the basic reproduction number (an epidemiological composite index of parameters and state variables) is globally identifiable even when individual model parameters are not, though we provide a counterexample demonstrating this does not hold universally.
ii. We demonstrate that when models are structurally nonidentifiable, identifiability can often be obtained by incorporating minimal noise-free data points in complementary observations. Specifically, we prove that even a single time-point observation from an additional data streams can be sufficient to achieve global identifiability—-a finding with important implications for surveillance design. This complements more standard approaches for achieving identifiability such as fixing parameters using external data sources (e.g., census data, laboratory experiments, clinical studies).

Additionally, we demonstrate that global identifiability of model parameters does not necessarily imply global identifiability of individual state variables. Furthermore, through examination of various models, particularly those with gamma-distributed latent periods and behavioral responses, we show that establishing general patterns of identifiability in classes of nonlinear models remains a nontrivial task.

We organize our findings by epidemiological modeling features that commonly arise in practice. We identify mathematical mechanisms causing non-identifiability (e.g., parameter products and sums) demonstrate strategies to achieve identifiability, and assess identifiability of epidemiologically relevant quantities like ℛ_0_ and state variables. Section 3.1 establishes the baseline SIR model and our methodology for assessing ℛ_0_ identifiability. Section 3.2 extends this to models with gamma-distributed disease progression times. Section 3.3 examines how partial observation of disease outcomes (specifically, infection fatality ratios) creates identifiability challenges. Section 3.4 addresses the transmission complexity arising from multiple transmission pathways and stratified populations. Section 3.5 investigates vector-borne disease dynamics with coupled host-vector populations. Finally, Section 3.5 explores how behavioral responses and nonlinear incidence functions affect the identifiability of both model parameters and the basic reproduction number.

### 3.1 SIR Model with Standard Incidence

The clasical SIR model with standard incidence provides a foundational example of structural non-identifiability arising from the quotient (or product) of two unknown parameters. Consider the model:

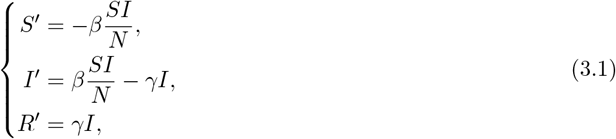

where *S, I*, and *R* represent susceptible, infectious, and recovered individuals, respectively, *β* is the transmission rate, *γ* is the recovery rate, and *N* = *S* + *I* + *R* is the total population.

#### Theorem

*Consider the SIR model* (3.1) *under observation of incidence (βSI/N) with unknown population size N. Then:*

i. *The parameter γ is globally identifiable, β and N are nonidentifiable; however, the parameter combination β/N is globally identifiable*.
ii. *The state variables S*(*t*) *and I*(*t*) *are globally identifiable, while R*(*t*) *is nonidentifiable*.
iii. *If the number of recovered individuals R*(*t*) *is known at a single time point, then the model becomes globally identifiable*.

The proof of Theorem 3.1 is given in Supplementary S1. Theorem 3.1 has several important implications.

First, Theorem 3.1(i) implies that the model cannot distinguish among a continuous spectrum of scenarios ranging from high transmission rates in large populations to low transmission rates in small populations, all yielding the same ratio *β/N* (see Figure 1(b) for visualization). Moreover, Theorem 3.1(ii) shows that *S* and *I* are identifiable while *R* is not (see the second row of Figure 1 for visualization). Therefore, an increase in *N* necessarily entails an increase in *R* (since *N* = *S* + *I* + *R* with *S* and *I* fixed). When *N* is large with correspondingly large *R*, a larger *β* is needed to penetrate the smaller susceptible fraction *S/N* and maintain the same transmission dynamic in the *S* and *I* compartments.

Second, although the model is structurally nonidentifiable, Theorem 3.1(ii) shows that some epidemiologically significant quantities such as *S* and *I* remain identifiable. Moreover, the effective reproduction number, given by 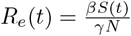, is identifiable since it depends only on the product of identifiable quantities: *γ, N/β*, and *S*. This identifiability of *R*_*e*_(*t*) is visually depicted in Figure 1(c) where, under the observation of incidence, the estimated effective reproduction number (grey curves) is indistinguishable from true *R*_*e*_(*t*) (green dashed curve).

**Figure 1.**
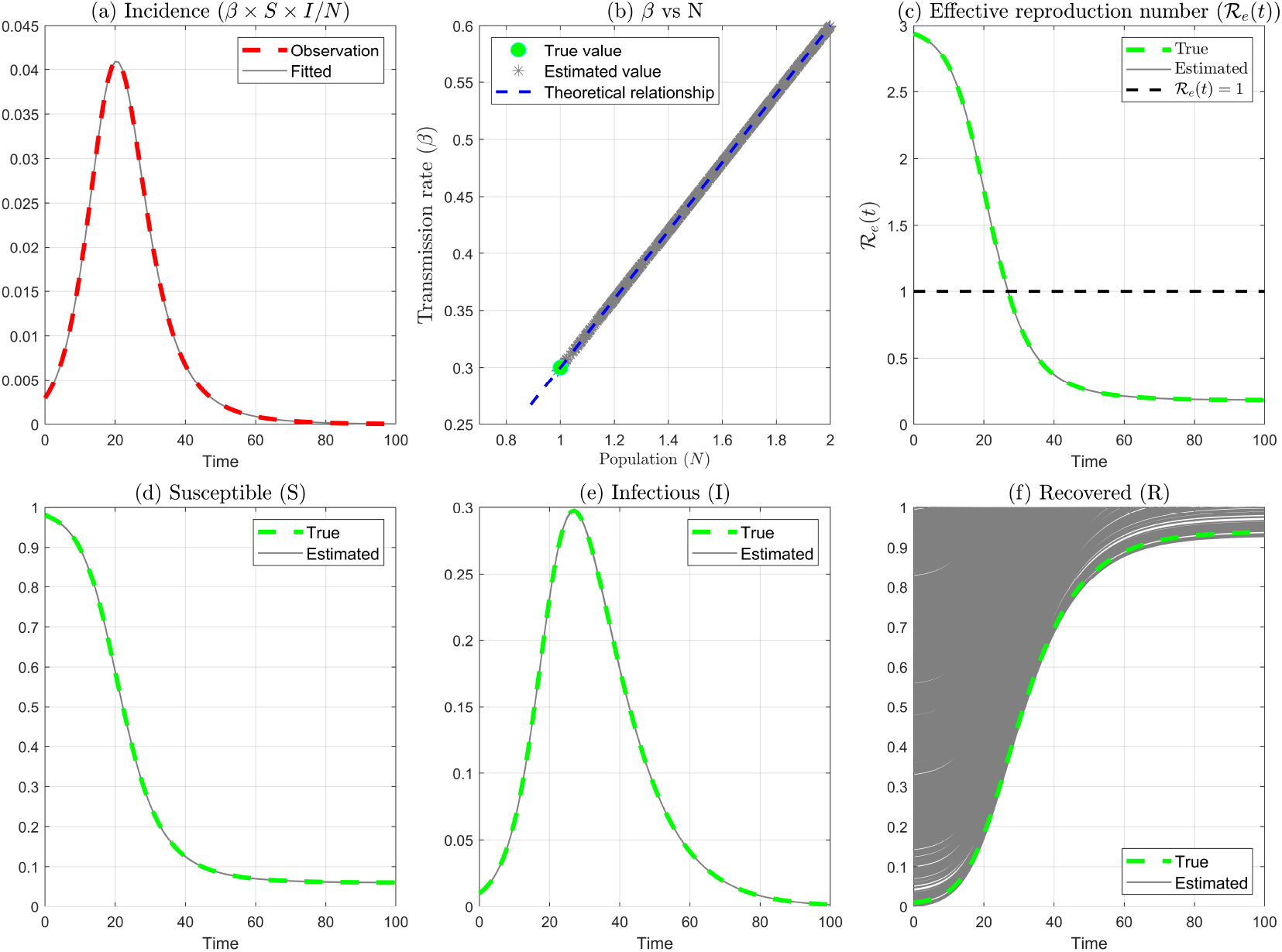
Identifiability issues in the SIR model (3.1) when simultaneously estimating transmission rate (*β*) and population size (*N*) from noise-free incidence data, demonstrated using multi-start optimization. **(a)** The fitted curves are visually indistinguishable from the true incidence data. **(b)** Estimated parameter pairs (*β, N*) shown as grey dots and and the true value (green dot) lie on the theoretical curve *β/N* = constant (blue dashed line). **(c)** The effective reproduction number *R*_*e*_(*t*) remains identifiable despite parameter non-identifiability, with all estimates (grey curves) matching the true trajectory (green dashed curve). **(d)-(f)** Susceptible and infectious populations are identifiable, while the recovered population is not. Data generation parameters: *β* = 0.3 day^−1^, *γ* = 0.1 day^−1^, *N* = 1, *I*(0) = *R*(0) = 0.01*N, S*(0) = *N* − (*I*(0) + *R*(0)). For simplicity, *γ* is fixed while *β, S*(0), *I*(0), and *R*(0) are estimated with *N* = *S*(0) + *I*(0) + *R*(0). Axes are truncated at *N* = 2 (panel b) and *R* = 1 (panel f) for visual clarity.

#### Remark 5

The effective reproduction number *R*_*e*_(*t*) of the SIR model (3.1), is structurally identifiable with respect to the observation of incidence, even though the model itself is structurally nonidentifiable. Moreover, ℛ_*e*_(*t* = 0) is globally identifiable. In the regime *S*(0) ≈ *N*, ℛ_*e*_(0) = ℛ_0_, which is also globally identifiable.

Since ℛ_0_ is also identifiable, so is herd immunity threshold 1 − 1*/*ℛ_0_. Moreover, because the state variable *I* is identifiable, key epidemic characteristics, including the peak time and maximum number of infected individuals, are also identifiable. Overall, these findings demonstrate that even when a model is structurally nonidentifiable under a given observation, epidemiologically important quantities may still be structurally identifiable.

Third, this structural nonidentifiable issue of the model (3.1) can be resolved if either the transmission rate *β* or the total population *N* is known independently. Since *β* varies spatially and temporally, the total population *N* —obtainable from census data or demographic projections—is often easier to determine. Alternatively, as stated in Theorem 3.1(iii), also knowing *R*(*t*) at a single time point is sufficient to achieve global identifiability. This is intuitively demonstrated in Figure 1(f), where fitted curves (gray bands) deviate substantially from the true dynamics (dashed green curve). However, if an additional constraint were imposed to force these gray bands to pass through even a single noise-free data point in *R*(*t*), it is intuitive that all the gray bands would collapse near the true dynamics depicted by the green curve.

#### Remark 6

The identifiability results of Theorem 3.1 hold for the SIR model (3.1) when the observation is active cases *I*(*t*) instead of incidence. In this case, the effective reproduction number is also globally identifiable (see Appendix A for details).

We note that for the remainder of this paper, unless otherwise stated, the initial population *N* (0), is assumed to be known.

### 3.2 Gamma-Distributed Latent and Infectious Periods

The standard SEIR model assumes exponentially distributed latent periods, which implies that the instantaneous rate of becoming infectious is constant and independent of time since infection. However, in reality, disease progression often follows more realistic distributions. A common approach to capture this biological realism is to subdivide compartments into sequential stages, which generates gamma-distributed residence times [11, 35]. This technique, known as the linear chain trick, allows modelers to represent more realistic disease progression dynamics while maintaining the computational tractability of ordinary differential equations. The resulting model with a Gamma-distributed latent period given by:

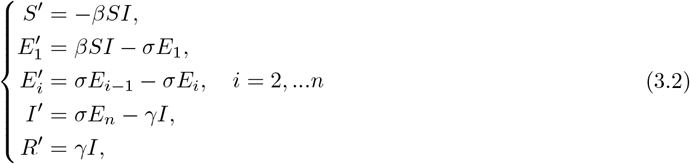

where *S, E*_*i*_ (with *i* = 1, …, *n*), *I*, and *R* represent susceptible, exposed (in stage *i*), infectious, and recovered individuals, respectively. The parameter *β* is the transmission rate, *σ* is the progression rate through each exposed stage, and *γ* is the recovery rate. The incubation period follows a gamma distribution with the shape parameter *n* and rate parameter *σ* (or scale parameter of 1*/σ*), yielding mean incubation period *n/σ*.

#### Theorem 3.2

*Consider the SE*_*n*_*IR model* (3.2) *under observation of incidence (σE*_*n*_*) with known initial population size* 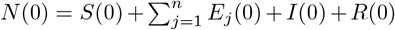. *Then the model is globally structurally identifiable, with all parameters and state variables being globally identifiable*.

The proof of Theorem 3.2 is given in Appendix B.

The basic reproduction number of model (3.2) is given by 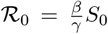. Since the parameters and initial conditions are globally identifiable, this quantity is globally identifiable as well.

We now examine a similar extension where the standard SIR model, which assumes exponentially distributed infectious periods, is generalized to incorporate gamma-distributed infectious periods. Specifically, infected individuals progress through *n* sequential infectious stages (each contributing to transmission) before recovering. The resulting model with a gamma-distributed infectious period is given by:

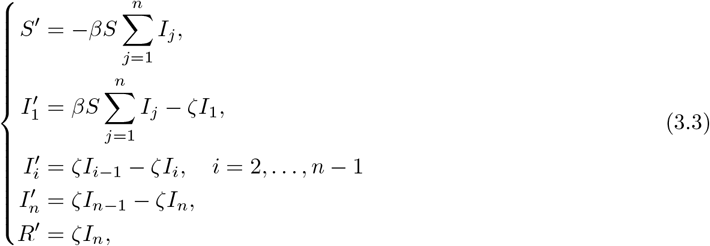

where *S, I*_*i*_ (with *i* = 1, …, *n*), and *R* represent susceptible, infectious (in stage *i*), and recovered individuals, respectively. The parameter *β* is the transmission rate and *ζ* is the progression rate through each infectious stage. The infectious period follows a gamma distribution with shape parameter *n* and rate parameter *ζ* (or scale parameter 1*/ζ*), yielding mean infectious period *n/ζ*.

The SI_*n*_R model (3.3) is globally structurally identifiable under the observation of incidence 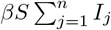 for *n* = 1, 2, 3, 4, 5 with known population size *N* (see Supplementary S2 for details). This implies that in this case, the basic reproduction number is also globally identifiable. Hence, we make the following unproven claim.

#### Conjecture 1

*The SI*_*n*_*R model* (3.3) *with gamma-distributed infectious periods is globally structurally identifiable under the observation of incidence* 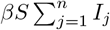 *for all n* ≥ 1 *and known population size* 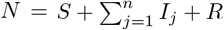.

### 3.3 Infection Fatality Ratio

The SIR model (3.1) can be extended to account for disease-induced mortality through two distinct parameterizations: a fractional approach where a fixed proportion of removals result in death, and an additive approach where recovery and mortality occur as independent competing processes.

#### Fractional Mortality Parameterization

Let 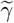 be the removal rate, with *ϕ*_*d*_ portion of removed individuals dying at a rate 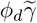 while the remaining (1 − *ϕ*_*d*_) recover at a rate 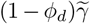. The resulting extension of the SIR model (3.1) is given by:

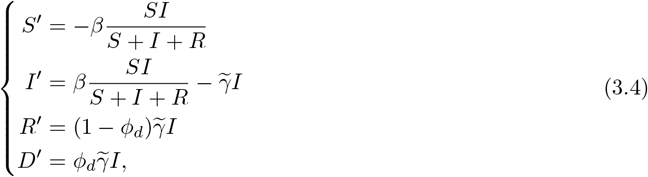

where *D* is the cumulative mortality.

##### Theorem 3.3

*Consider the SIRD model* (3.4) *under observation of new deaths* 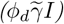 *with known initial population size N* (0) = *S*(0) + *I*(0) + *R*(0). *Then:*

i. *The model is structurally nonidentifiable. Specifically*, 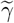 *is globally identifiable and so is the ratio β/ϕ*_*d*_, *while β and ϕ*_*d*_ *are nonidentifiable*.
ii. *Moreover, while the individual state variables S*(*t*), *I*(*t*), *and R*(*t*) *are nonidentifiable, the scaled quantities ϕ*_*d*_*S*(*t*) *and ϕ*_*d*_*I*(*t*) *are globally identifiable*.
iii. *The model becomes globally structurally identifiable if even a single noise-free data point from S*(*t*), *I*(*t*), *or R*(*t*) *is also observed*.

The proof of Theorem 3.3 is given in Supplementary S3.

Theorem 3.3 has important implications. First, the non-identifiability manifests not only as correlations between parameters but also as correlations between parameters and initial conditions. A continuum of scenarios—ranging from high transmission rates (*β*) with low infection fatality ratios (*ϕ*_*d*_) to low transmission rates with high infection fatality ratios—can generate identical mortality observations, all constrained to lie along the identifiable ratio *β/ϕ*_*d*_ (Figure 2(d) for visualization). Similarly, theoretical relationships exist between *ϕ*_*d*_ and the initial conditions *S*(0) and *I*(0) (Figure 2(e)–(f)), with estimated values lying on these theoretical curves. The relationship between *ϕ*_*d*_ and *R*(0) (Figure 2(g)) is constrained by *N* (0) = *S*(0)+*I*(0)+ *R*(0); this constraint structure is captured in Figure 2(c). These parameter–initial condition correlations would be missed by identifiability analyses that focus solely on model parameters, yet they are essential for understanding why complementary observations (including knowing initial conditions) can resolve non-identifiability.

**Figure 2.**
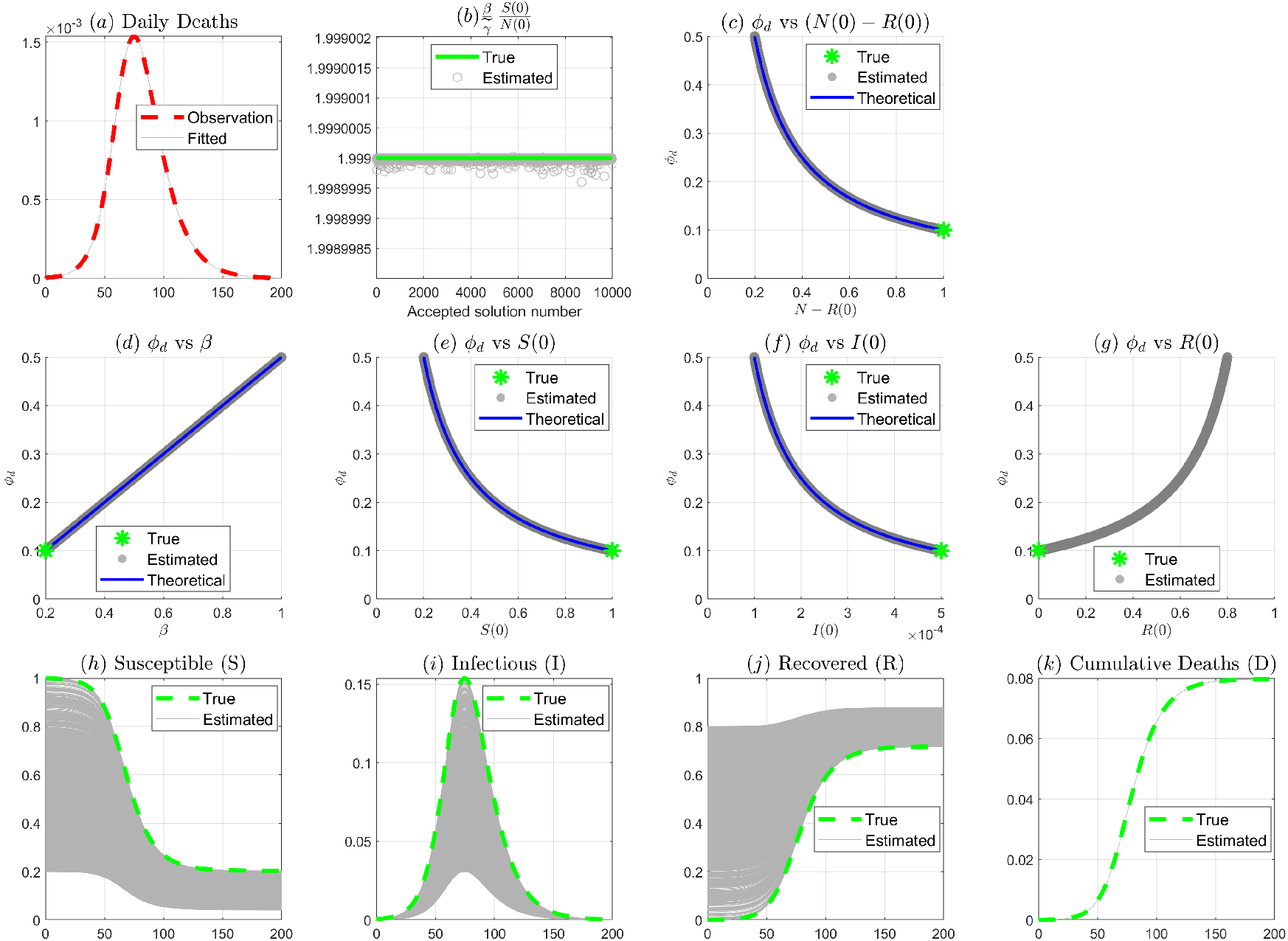
Visualization of identifiability issues when fitting the SIRD model (3.4) to daily deaths. **(a)** Model-fitted curves (gray lines) are visually indistinguishable from observed daily death data (red dashed). **(b)** The quantity, 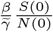 is identifiable across accepted solutions. **(c)** Parameter correlation between *ϕ*_*d*_ and (*N* − *R*(0)): estimated values lie on the theoretical curve (blue) and the true value (green star) satisfies this constraint. **(d)-(f)** Parameter correlations showing estimated values lie on theoretical curves alongside the true value: *ϕ*_*d*_ vs *β, ϕ*_*d*_ vs *S*(0), and *ϕ*_*d*_ vs *I*(0). **(g)** Scatter of *ϕ*_*d*_ vs *R*(0) values; due to the constraint *N* (0) = *S*(0) + *I*(0) + *R*(0), the relationship between *ϕ*_*d*_ and *R*(0) is determined by the constraints in (e) and (f), as depicted in (c).**(h)-(k)** Model trajectories: susceptible, infectious, recovered, and cumulative deaths. Gray curves show fitted solutions; green dashed lines show true dynamics. The parameters used to generate the synthetic data are: *β* = 0.2, *γ* = 0.1, *ϕ*_*d*_ = 0.1, *N* = 1, *I*(0) = 5 *×* 10^−4^, *R*(0) = 0, and *S*(0) = 1 − *I*(0). For simplicity, we fix *γ* (which is globally structurally identifiable) in the data fitting process, while the remaining parameters and initial conditions are fitted with the constraint *N* (0) = *S*(0) + *I*(0) + *R*(0).

Second, although the SIRD model (3.4) is structurally nonidentifiable under observation of new deaths alone, the basic reproduction number of the model (3.4), defined as 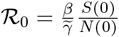, remains globally identifiable. Since *N* (0) is assumed to be known and 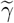 is identifiable, ℛ_0_ is identifiable if and only if *βS*(0) is identifiable. From Theorem 3.3(ii), the product *βS*(*t*) is identifiable for all *t* (specifically, *βS*(*t*) = (*β/ϕ*_*d*_) · (*ϕ*_*d*_*S*(*t*)) where both factors are identifiable). Therefore, *βS*(0) is identifiable, and consequently ℛ_0_ is globally structurally identifiable (see Figure 2(b) for visual illustration). This highlights that epidemiologically significant composite quantities can be identifiable even when individual parameters are not.

##### Remark 7

The basic reproduction number of the SIRD model (3.4) is globally structurally identifiable under observation of new mortality, although the model itself is structurally nonidentifiable.

Third, Theorem 3.3(iii) shows that incorporating even a single noise-free data point from *S*(*t*) or *I*(*t*) resolves all identifiability issues. The mechanism can be understood from Theorem 3.3(ii): although individual state variables are not identifiable, the scaled quantities *ϕ*_*d*_*S*(*t*) and *ϕ*_*d*_*I*(*t*) are identifiable. A single observation from *S*(*t*) or *I*(*t*) determines the scaling factor *ϕ*_*d*_ which, combined with the identifiable ratio *β/ϕ*_*d*_ from Theorem 3.3(i), identifies *β*, consequently leading to identifiability of all state variables, and hence global identifiability of the entire model. Figure 2(h)–(k) illustrate this intuitively: the gray trajectory bundles in *S*(*t*), *I*(*t*), and *R*(*t*) would collapse near the true dynamics (green dashed) if constrained to pass through even a single observed point in *S*(*t*) or *I*(*t*).

#### Additive Mortality Parameterization

The fractional mortality structure in model (3.4) can be reparameterized by decomposing the removal rate 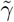 into two distinct rates: recovery rate *γ* and mortality rate *δ*, i.e., 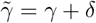. Setting 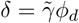 and 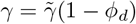 yields a common alternative formulation where recovery and death occur as independent competing processes:

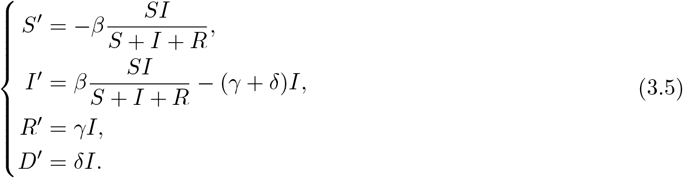

In model (3.5) the infection fatality ratio is given by *δ/*(*γ* + *δ*), which is equivalent to *ϕ*_*d*_ in model (3.4).

Under observation of new deaths (*δI*), model (3.5) is also structurally nonidentifiable. Since model (3.5) is a reparameterization of model (3.4) with 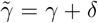, the identifiability of 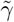 from Theorem 3.3 immediately implies that *γ* +*δ* is globally identifiable. Moreover, the relationship 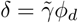 combined with the identifiability of *β/ϕ*_*d*_ yields the identifiability of *β/δ*. However, *β, γ*, and *δ* are nonidentifiable. This non-identifiability arises from the observation’s inability to decompose the total removal rate into its two additive components (*γ* and *δ*), illustrating how additive rate structures can create identifiability challenges. This contrasts with model (3.4), where non-identifiability stems from a multiplicative factor.

### 3.4 Multiple Transmission Pathways

In this section, we consider two distinct examples of multiple transmission modes: a cholera model with both human-to-human and waterborne transmission, and a stratified population model with aggregated observations.

#### Cholera model

Consider the following simplified version of the cholera model presented in [23]:

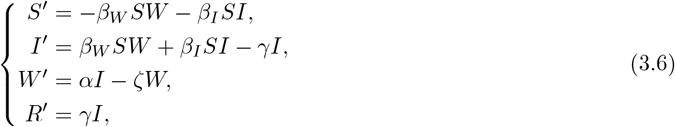

where it is assumed that susceptible individuals (S) are infected by infectious individuals (I) at a rate *β*_*I*_ and by the concentration of the pathogen in water (W) at a rate *β*_*W*_. Infected individuals are assumed to shed pathogens in the water at a rate *α* and pathogens decay in the water at a rate *ζ*. We analyze identifiability under two related observations: pathogen concentration in the water (*W*) and new pathogen inflow (*αI*).

##### Theorem 3.4

*Consider the cholera model* (3.6) *under observation of pathogen concentration in the water (W) with known initial population size N* (0) = *S*(0) + *I*(0) + *R*(0). *Then:*

i. *None of the model parameters are globally identifiable. However, γζ, γ* + *ζ, α/β*_*I*_, *and* (*β*_*W*_ *α* + *β*_*I*_*ζ*)*/β*_*I*_ *are globally identifiable. Parameters ζ, γ and β*_*W*_ *are locally identifiable*.
ii. *Moreover, while the individual state variables S*(*t*), *I*(*t*), *and R*(*t*) *are nonidentifiable, the following are globally identifiable: β*_*I*_*S*(*t*) *and β*_*I*_*I*(*t*) + *β*_*W*_ *W* (*t*).
iii. *The model becomes globally structurally identifiable if one noise-free data point from each of two distinct epidemiological variables among S*(*t*), *I*(*t*), *and R*(*t*) *is also available*.

##### Theorem 3.5

*Consider the cholera model* (3.6) *under observation of new pathogen inflow (αI) with known initial population size N* (0) = *S*(0) + *I*(0) + *R*(0). *Then:*

i. *Parameters ζ, γ, and β*_*W*_ *are globally identifiable, alongside the parameter combination α/β*_*I*_, *but α and β*_*I*_ *is nonidentifiable*.
ii. *Moreover, the state variable S*(*t*), *I*(*t*), *and R*(*t*) *are nonidentifiable. However, the following scaled quantities are globally identifiable: β*_*I*_*S*(*t*) *and β*_*I*_*I*(*t*).
iii. *The model becomes globally structurally identifiable if even a single noise-free data point from S*(*t*), *I*(*t*), *or R*(*t*) *is also available*.

The proofs of Theorem 3.4 and Theorem 3.5 are given in Supplementary Sections S4 and S5, respectively. Theorem 3.4 and Theorem 3.5 reveal several important insights about identifiability in models with multiple transmission pathways. First, the choice of observation—even when measuring the same underlying environmental reservoir—fundamentally alters which parameters become identifiable. Observing pathogen concentration (*W*) directly yields identifiability of the product *γζ* and sum *γ* + *ζ*, but not the individual recovery and pathogen decay rates. In contrast, observing the rate of pathogen introduction (*αI*) makes both *γ* and *ζ* individually identifiable.

Second, despite stark differences in individual parameter identifiability between the two observation scenarios, the basic reproduction number 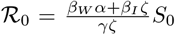 remains globally structurally identifiable under both observations, as it decomposes into a product of globally identifiable quantities. Under observation of *W* (*t*), ℛ_0_ decomposes as

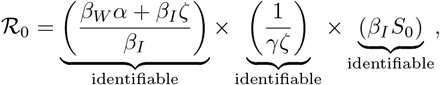

while under observation of *αI*, it decomposes as

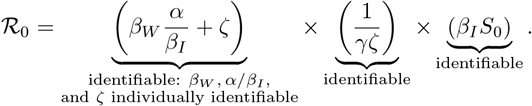

##### Remark 8

The basic reproduction number of the cholera model (3.6) is globally structurally identifiable under observation of either pathogen concentration or new pathogen inflow, even though the model itself is structurally nonidentifiable.

Third, these theorems reveal a crucial asymmetry in data requirements for achieving global identifiability. When observing pathogen concentration *W* (*t*), restoring full identifiability requires two complementary data points—one each from two distinct compartments among *S*(*t*), *I*(*t*), and *R*(*t*). However, when observing pathogen inflow *αI*, even a single data point from any one of these compartments suffices. This difference has important implications for surveillance strategy. From a practical surveillance perspective, this suggests that measuring pathogen inflow rates rather than standing concentrations reduces data requirements for parameter estimation—requiring only a single time point from one data stream rather than from two.

The mechanism underlying this asymmetry can be understood from the structure of identifiability results: under observation of *W* (*t*), Theorem 3.4(ii) shows that scaled quantities *β*_*I*_*S*(*t*) and *β*_*I*_*I*(*t*) + *β*_*W*_ *W* (*t*) are identifiable, and two complementary observations are needed to determine both scaling factors *β*_*I*_ and *β*_*W*_, which combined with the identifiable parameter combinations from Theorem 3.4(i) (*γζ, γ* + *ζ*, and *α/β*_*I*_), identify all parameters. In contrast, under observation of *αI*, Theorem 3.5(ii) shows that *β*_*I*_*S*(*t*), *β*_*I*_*I*(*t*), and *β*_*I*_*R*(*t*) are identifiable, and a single complementary observation determines the scaling factor *β*_*I*_, which combined with the individually identifiable parameters from Theorem 3.5(i) (*ζ, γ, β*_*W*_, and *α/β*_*I*_), identifies all parameters.

#### Aggregated Observations

Consider the following model that stratifies the population into two groups (e.g., age groups, geographic regions, or risk categories) but makes a naive simplifying assumption that both groups share identical parameters. This leads to the following model that artificially stratifies a homogeneous population into two groups:

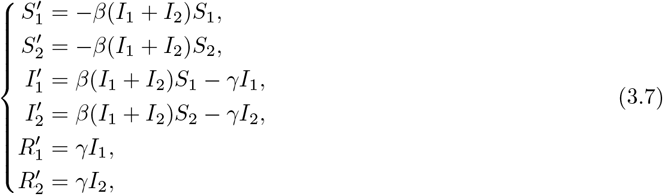

where *S*_*i*_, *I*_*i*_, and *R*_*i*_ represent susceptible, infectious, and recovered individuals in subpopulation *i* for *i* = 1, 2, with both subpopulations having identical transmission rate *β* and recovery rate *γ*.

##### Theorem 3.6

*Consider the stratified SIR model* (3.7) *under observation of total active infectious individuals (I*_1_ + *I*_2_*) with known initial population sizes of each group (i.e*., *N*_*i*_(0) = *S*_*i*_(0) + *I*_*i*_(0) + *R*_*i*_(0) *with i* = 1, 2*). Then:*

i. *Both parameters β and γ are globally identifiable*.
ii. *The individual state variables S*_1_(*t*), *S*_2_(*t*), *I*_1_(*t*), *and I*_2_(*t*) *are nonidentifiable; however, the aggregate quantities S*_1_(*t*) + *S*_2_(*t*) *and I*_1_(*t*) + *I*_2_(*t*) *are globally identifiable*.

The proof of Theorem 3.6 is given in Supplementary S6.

Theorem 3.6 illustrates that global identifiability of all model parameters does not guarantee global identifiability of state variables. Furthermore, it showcases the difficulty in extracting individual subpopulation dynamics from aggregate observations. For example, observing 100 total infectious individuals provides no information about burden distribution: whether equally split (50 each) or concentrated (90 in one, 10 in the other). Importantly, the model (3.7) can be rewritten in terms of aggregate variables *S* := *S*_1_ + *S*_2_, *I* := *I*_1_ + *I*_2_, *R* := *R*_1_ + *R*_2_, yielding a standard SIR model. This exact reduction from a stratified to an aggregate system is an example of *lumping* [10, 20, 39], a technique that groups states or variables to reduce system dimension while preserving model dynamics, and represents a broader class of models where parameter identifiability does not imply state identifiability.

### 3.5 Host-Vector Dynamics

We consider the following form of Ross–Macdonald model [29] that divides the constant human population (*H*) into infectious (*I*_*h*_) and susceptible (*H* − *I*_*h*_) compartments, and the constant mosquito population (*V*) into infectious (*I*_*v*_) and susceptible (*V* − *I*_*v*_) compartments:

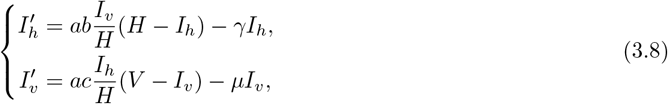

where *a* is the biting rate, *b* and *c* are transmission probabilities from mosquito-to-human and human-to-mosquito respectively, *γ* is the human recovery rate, *µ* is the mosquito death rate.

#### Theorem 3.7

*Consider the Ross-Macdonald model* (3.8) *under observation of incidence* 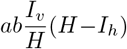 *where the constant human population H and the constant mosquito population V are assumed to be unknown. Then:*

i. *Parameters γ, µ, and H are globally identifiable, and so are the parameter combinations ac and abV*.
ii. *Moreoever, state variable I*_*h*_ *is globally identifiable while I*_*v*_ *is nonidentifiable. However*, 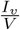 *is globally identifiable*.

The proof of Theorem 3.7 is given in Supplementary S7.

Theorem 3.7 has the following identifiability implications. It reveals that the Ross-Macdonald model (3.8) becomes structurally identifiable when either (1) transmission probability from mosquito-to-human (*b*), (2) mosquito population (*V*), or (3) a single data point in *I*_*v*_(*t*) is known along with one of the following:

- biting rate (*a*), or
- transmission probability from human-to-mosquito (*c*)

Estimating mosquito population (*V*) or single observation of infected mosquitoes (*I*_*v*_) requires labor-intensive field surveillance such as trapping, mark-release-recapture studies, or oviposition monitoring, while measuring biting rate (*a*) demands resource-intensive human landing catches or feeding assays. In contrast, transmission probabilities (*b* and *c*) may be determined more readily through controlled laboratory membrane feeding experiments and blood meal analyses. Consequently, fixing *b* and *c* based on laboratory measurements is likely a more practical approach to achieving structural identifiability.

The basic reproduction number of model (3.8), given by 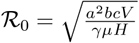 [29], is globally structurally identifiable as the quantity under the square root decomposes into quotients and products of globally identifiable quantities. Specifically, the denominator *γµH* is a product of globally identifiable parameters, while the numerator *a*^2^*bcV* can be expressed as a product of (*ac*) and (*abV*), both of which are also globally identifiable per Theorem 3.7(i).

#### Remark 9

The basic reproduction number of the Ross-Macdonald model (3.8) is globally identifiable under the observation of incidence, even though the model itself is structurally nonidentifiable.

### 3.6 Behavioral Response and Nonlinear Incidence

In the various model structures considered thus far, the basic reproduction number has been shown to be globally identifiable, even when a model was not globally identifiable with respect to an observation. However, this property does not hold universally, as we illustrate using a behavioral-epidemiological model.

Several studies have incorporated behavior change into epidemiological models by considering feedback between disease state and transmission, such as the number of infectious individuals or reported deaths [32, 41, 42]. We examine a simple behavioral extension of the SIR model where individuals reduce contact rates as infections increase. Specifically, we assume that the transmission rate depends on the population of infectious individuals (*I*) and is given by *β*(*t*) = *β*_0_*/*(1 + *αI*)^*k*^, where *β*_0_ is the baseline transmission rate in the absence of behavioral response and *α* measures the strength of the behavioral response to infection prevalence and *k* ≥ 1 is the saturation exponent. The resulting behavioral response model is given by:

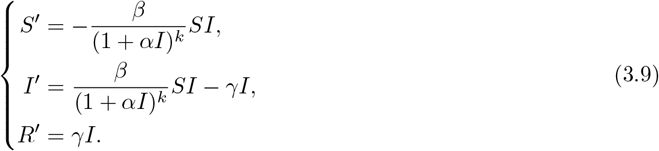

#### Theorem 3.8

*Consider the behavioral response model* (3.9) *with saturation exponent k* = 1 *under observation of active cases I*(*t*) *with total initial population N* (0) = *S*(0) + *I*(0) + *R*(0) *known. Then:*

i. *The parameter α is globally identifiable while β and γ are only locally identifiable. Moreover, the following parameter combinations are globally identifiable: βγ and αγ* + *β*.
ii. *The state variables S*(*t*) *and R*(*t*) *are only locally identifiable, while the observation I*(*t*), *is globally identifiable; although β*(*S* + *I*) − *γ is globally identifiable*.
iii. *The model becomes globally structurally identifiable if even a single noise-free data point from S*(*t*) *or R*(*t*) *is also available*.

The proof of Theorem 3.8 is given in Supplementary S8.

Theorem 3.8 has several implications. First, since *βγ* is globally identifiable, fixing the recovery rate *γ* makes all parameters of the model (3.8) globally identifiable (see Theorem 3.8(i)). Furthermore, since *β*(*S* +*I*) − *γ* is globally identifiable (see Theorem 3.8(ii)), the global identifiability of parameters implies global identifiability of the state variable *S*, and consequently *R* = *N* − *S* − *I* is also identifiable with the known population size. Hence, fixing parameter *γ* makes the model (3.8) globally identifiable.

Second, in contrast to the previous models where ℛ_0_ was globally identifiable even when the model was not, the basic reproduction number of behavioral model (3.9), given by 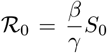, is only locally identifiable. Since *β, γ*, and *S* are locally identifiable, ℛ_0_ is also locally identifiable. However, combinations of locally identifiable quantities can sometimes yield globally identifiable functions (see Remark 3), so we now demonstrate explicitly that ℛ_0_ for behavioral model (3.9) is *not globally identifiable*.

Consider a trajectory (*S*^∗^(*t*), *I*^∗^(*t*), *R*^∗^(*t*)) of (3.9) with parameter values (*α*^∗^, *β*^∗^, *γ*^∗^). We can obtain another trajectory (*S*^*†*^(*t*), *I*^*†*^(*t*), *R*^*†*^(*t*)) and parameter values (*α*^*†*^, *β*^*†*^, *γ*^*†*^) yielding the same observation by equating the identifiable quantities from the theorem

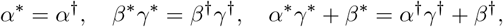

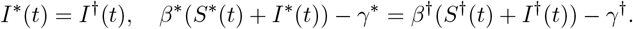

Solving the above equations, we find

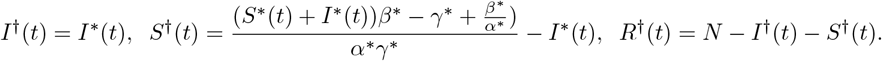

and *α*^*†*^ = *α*^∗^, *β*^*†*^ = *α*^∗^*γ*^∗^, and 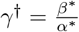. Then one can verify by a direct computation that (*S*^*†*^(*t*), *I*^*†*^(*t*), *R*^*†*^(*t*)) is indeed a solution of of (3.9) with parameter values (*α*^*†*^, *β*^*†*^, *γ*^*†*^). Since *I*^∗^(*t*) = *I*^*†*^(*t*), these two solutions have the same output data. However, they have different basic reproduction numbers:

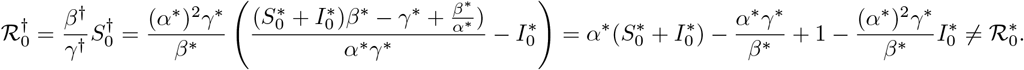

#### Remark 10

Under observation of active cases *I*(*t*), the basic reproduction number of the behavioral model (3.9) with *k* = 1 is locally (but not globally) structurally identifiable.

Third, model (3.9) is globally identifiable if even a single noise-free data point from *S*(*t*) or *R*(*t*) is also observed.

It turns out that the behavioral model (3.9) with *k* = 2, 3, 4 is globally structurally identifiable under the observation of active cases *I*(*t*) (see Supplementary Section S9 for details and additional evidence). Hence, we make the following unproven statement.

#### Conjecture 2

*The behavioral model* (3.9) *with k* ≥ 2 *is globally structurally identifiable under the observation of active cases I*(*t*) *with total initial population N* (0) = *S*(0) + *I*(0) + *R*(0) *known*.

### 4 Discussion

This study makes two key contributions to the analysis of structural identifiability in epidemiological models. First, we present a methodology to assess the structural identifiability of epidemiologically relevant quantities of interest, such as the basic reproduction number ℛ_0_. We demonstrate that models that are structurally nonidentifiable can still yield structurally identifiable quantities of interest that are epidemiologically useful for decision-making. Second, we show that strategic incorporation of complementary data–often requiring as little as a single time-point observation–can resolve identifiability issues.

A noteworthy finding is the remarkable robustness of the basic reproduction number identifiability across vastly different model structures. Specifically, for nearly all models considered in this study—including cholera transmission with multiple pathways and host-vector dynamics—the basic reproduction number (ℛ_0_) is globally structurally identifiable even when the model itself is not. The sole exception is the behavioral response model with a saturating incidence (*k* = 1), where ℛ_0_ exhibits only local identifiability. Despite these models having vastly different transmission mechanisms and observation (e.g., direct human-to-human transmission to water-mediated pathways to vector-borne dynamics) and different observations (e.g., active cases, incidence, and pathogen concentration in water), ℛ_0_ maintains this robust identifiability property. This structural robustness complements findings from practical identifiability studies—which assess whether parameters can be reliably estimated from finite, noisy data—showing that ℛ_0_ can often be estimated more precisely than individual parameters [30, 48]. Mathematically, ℛ_0_ becomes identifiable because this composite quantity can be decomposed as a combination of various identifiable terms. However, whether such a structure arises from deeper theoretical principles or is coincidental across the models examined in this manuscript remains unclear. Understanding what mathematical features guarantee ℛ_0_ identifiability, and characterizing the conditions under which this property fails, represents an important open theoretical question [13, 31].

We present and utilize a systematic methodology to resolve structural identifiability issues through the strategic incorporation of a minimal number of data points in complementary observations. Our analysis reveals that when a model is structurally nonidentifiable under continuous observation of a data stream, incorporating even a single noise-free measurement from a complementary state variable can achieve global identifiability. For instance, in the SIRD model (3.4) where mortality data alone cannot distinguish between high transmission with low case-fatality versus low transmission with high case-fatality, a single observation from *S*(*t*), *I*(*t*), or *R*(*t*) resolves this degeneracy and renders the model globally identifiable (Theorem 3.3). The cholera model (3.6) demonstrates that data requirements depend on observation type: measuring pathogen inflow requires only one complementary observation (Theorem 3.5), while measuring pathogen concentration requires two from distinct compartments (Theorem 3.4). Overall, these findings have profound implications for surveillance system design. Rather than asking “how much data do we need in our data streams?”, the critical question becomes “which additional complementary measurements bring new information about disease dynamics?” This shift in perspective prioritizes the strategic collection of high-quality sparse data from multiple complementary streams over increasing measurement frequency within a single data stream. Such an approach is particularly valuable in resource-constrained settings where comprehensive multi-stream surveillance is infeasible. Under ideal conditions (including perfect test sensitivity and specificity, representative sampling, and persistent antibody detection), a serological measurement represents a single observation of the susceptible population (i.e., a data point in *S*(*t*)) [43], suggesting that even periodic cross-sectional serological surveys, despite their sparsity, can provide the complementary information necessary to resolve fundamental identifiability issues.

Structural identifiability issues, once identified through rigorous analysis, may appear obvious in retrospect, particularly when visualized with state observability results in hand. However, this retrospective clarity should not obscure the genuine difficulty of recognizing these patterns prospectively, especially as the model complexity increases. Our analysis reveals that non-identifiability often arises from specific mathematical structures across diverse epidemiological models: parameter quotients (e.g., *β/N* in model (3.1)), products (e.g., *ϕ*_*d*_*γ* in model (3.4)), and sums (e.g., *γ* + *δ* in model (3.5)) emerge as identifiable combinations when their individual components are not. The behavioral response model yields an even more complex globally identifiable combination *β*(*S* + *I*) − *γ* (Theorem 3.8), which mixes parameters with state variables in a non-trivial way. Critically, these patterns arise not solely from model structure but from the interaction between model structure and available observations. A model may be structurally nonidentifiable under one observation yet globally identifiable under another, even when both observations measure epidemiologically related quantities [9]. Furthermore, even within structurally similar model families, identifiability patterns can be surprisingly subtle. The behavioral response model (3.9) illustrates this difficulty: with saturation exponent *k* = 1, the model exhibits only local identifiability, while for *k* ≥ 2 it appears globally identifiable. Such sensitivity to structural details, even within a single model class, highlights why establishing general identifiability principles remains challenging for nonlinear epidemiological models. While systematic identifiability characterizations exist for some infinite series of linear dynamical systems (e.g.[7, 8, 24, 37]), extending such results to nonlinear models represents a significantly more difficult task. Our proof establishing global identifiability for the *SE*_*n*_*IR* model with gamma-distributed latent periods (Theorem 3.2) represents progress in this direction, yet the complexity of this proof underscores the fundamental challenges in deriving general identifiability results for nonlinear epidemic models.

Due to model complexity and the inherent limitations of surveillance systems (where, for example, detected cases capture only a fraction of true infections), structural non-identifiability is often present in mathematical epidemiology and mathematical biology more broadly. However, this need not be problematic. As demonstrated throughout this study, structurally nonidentifiable models can still produce identifiable composite quantities (such as the basic reproduction number) and observable state variables that are directly relevant for public health decision-making. This observation connects naturally to the adequacy-for-purpose framework, which evaluates models based on their fitness for specific intended uses rather than general accuracy [5, 6, 44]. A structurally nonidentifiable model-observation pairing may be adequate for purpose if the goal is to determine the basic reproduction number, a threshold quantity particularly valuable in early outbreak stages for assessing whether the disease will spread and what intervention intensity is required to curtail transmission. The same model-observation pairing may prove inadequate for the purpose if the goal shifts to estimating when the epidemic will peak or the maximum case burden during an outbreak. Hence, we propose reframing the identifiability question from “is the model-observation paring structurally identifiable?” to “are the quantities that matter for the decision at hand structurally identifiable?”

## Data Availability

Code for all analyses and figures is available at https://github.com/Binod-Pant/structural-identifiability-complementary.

## Acknowledgments

GP was supported by the French ANR-22-CE48-0008 OCCAM project. OS acknowledges the support from the Virginia Tech Center for the Mathematics of Biosystems (VTCMB-033500). BP was supported by cooperative agreement CDC-RFA-FT-23-0069 from the CDC’s Center for Forecasting and Outbreak Analytics. The views expressed in written in this publication do not necessarily reflect the official policies of the Department of Health and Human Services/Centers for Disease Control and Prevention.

## Appendix A Full identifiability analysis of the SIR model with the active cases observation

In this section, we demonstrate the workflow for analyzing structural identifiability described in Section 2.2 using the following version of Theorem 3.1.

### Theorem A.1

*Consider the SIR model* (A.1) *with unknown parameters β, γ, and N, and the number of active cases, given by I*(*t*), *as the observation*.

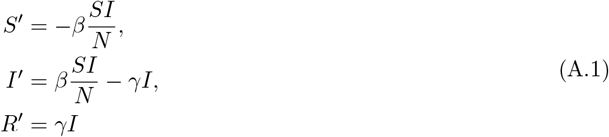

*Then:*

i. *Parameter β and N are nonidentifiable, however γ and β/N are globally identifiable;*
ii. *The state variables S*(*t*) *and I*(*t*) *are globally identifiable, while R*(*t*) *is nonidentifiable;*
iii. *If the number of recovered individuals R*(*t*) *is known at a single time point, then the model becomes globally identifiable*.

*Proof*. We will prove the three statements of the theorem while highlighting the steps of identifiability analysis from Section 2.2. The general correspondence (which is also true for the proofs in the Supplementary Materials) is the following:

i. identifiability analysis of parameters is accomplished by **(Step 1)**-**(Step 3)**;
ii. identifiability of states is then assessed in **(Step 4)**;
iii. the last part of the theorem is not a part of the four-step procedure outline in Section 2.2 but relies on the results obtained there.

We start by proving part (i) of the theorem.

**(Step 1)**: eliminating the states and obtaining the minimal order relation for the observation *y* = *I* and the parameters. Plugging *y* = *I* into second equation of (A.1), we have

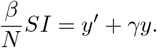

Plugging this into the first equation of (A.1), we obtain

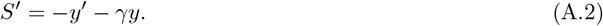

On the other hand, the second equation of (A.1) implies 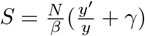, so by differentiating we get

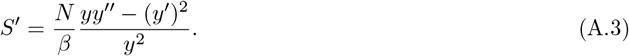

Equating the right-hand sides of (A.2) and (A.3), we get

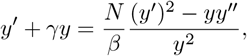

which can be rewritten in polynomial form as

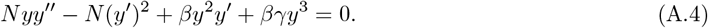

This is an irreducible second order differential equation for *y* and the algebraic independence of *y* = *I* and 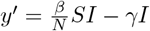 implies that this is the lowest possible order.

**(Step 2)**: checking if the coefficients of (A.4) give a complete set of identifiable combinations. We now look at the input-output equation A.4 as a polynomial in *y* and its derivatives and consider its coefficients:

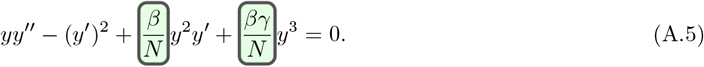

Note that we have normalized the coefficients so that at least one of them is identity, this is important to make the equation being uniquely defined, not up to a scaling. Then, by [40, Theorem 1]^1^, every identifiable combination of parameters can be expressed via these two coefficients. Thus, in order to establish that these coefficients give a complete set of identifiable combinations, it is sufficient to establish their identifiability. [40, Lemma 1]^2^ shows that this can be done by proving the the Wronskian of the corresponding monomials, *y*^2^*y*^*′*^ and *y*^3^ is nonsingular. Before verifying this, let us give a rationale why the nonsingularity of the Wronskian indeed implies identifiability of the coefficients.

Differentiating (A.5) gives:

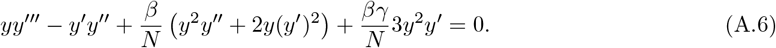

Equations (A.5) and (A.6) can be written as a linear system with respect to 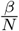 and 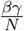:

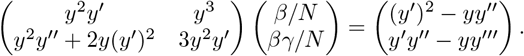

The matrix of this system is the Wronskian of *y*^2^*y*^*′*^ and *y*^3^. Therefore, if the Wronskian is nonsingular, the values of 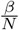 and 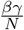 can be expressed in terms of the observable and its derivatives using the Cramer’s rule. In order to demonstrate the nonsingularity, we evaluate the determinant

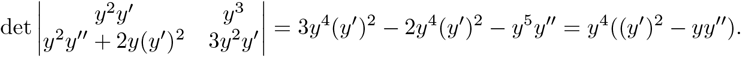

Expressing the last expression in terms of *S, I*, and parameters, we obtain:

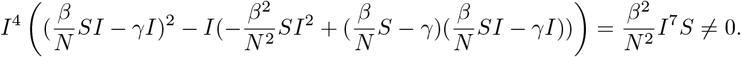

**(Step 3)**: establishing identifiability of parameters and computing identifiable combinations. In this particular case, it is easy to see that 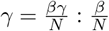 is a quotient of globally identifiable quantities, so *γ* is globally identifiable as well as 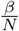. On the other hand, *β* and *N* are nonidentifiable since any scaling transformation *β*^∗^ = *λβ, N*^∗^ = *λN* with nonzero *λ* yields another parameter value giving he same IO-equation in arbitrary small neighborhood of the original parameter values. This concludes the proof of part (i) of the theorem.

We now continue with proving part (ii) of the theorem.

**(Step 4)**: analyze the Lie derivatives of the outputs to establish the identifiability of the states. The state *I* is clearly identifiable since it is observed. In order to establish identifiability of *S*, we consider the first derivative of *y*:

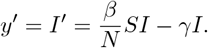

Solving with respect to *S* gives us

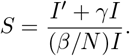

Since the right-hand side is an arithmetical expression in identifiable quantities *I, I*^*′*^, *γ, β/N*, the left-hand side is identifiable. Finally, we note that *R* does not appear in any of the equations related to output and cannot be identified from the relation *N* = *S* + *I* + *R* since *N* is not identifiable itself. Thus, *R* is not identifiable. This finished the proof of part (ii) of the theorem.

We conclude with proving part (iii) of the theorem. If *R* is known at some point and since *S* and *I* are globally identifiable, in the generic case (i.e., on the open dense subset ensuring uniqueness of *I* and *S*), the sum *S* + *I* + *R* = *N* is known at this point. Hence, *N* is identifiable. Together with identifiability of 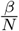 this implies identifiability of *β*. Finally, since *S, I, N* are identifiable, so is *R* = *N* − *S* − *I*.

## Appendix B Proof of Theorem 3.2

We will consider the following series of models Σ_*n*_:

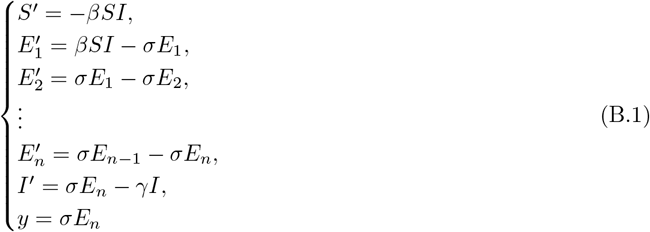

### Theorem B.1

(Theorem 3.2). *All the parameters and states in* (B.1) *are globally identifiable*.

We will start by performing a linear change of variables:

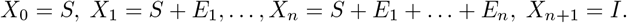

In these coordinates, the system looks as follows:

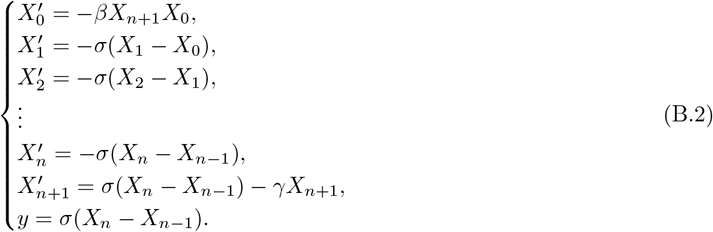

In order to be able to use input-output equations to establish identifiability, we will prove that the model does not admit rational first integrals. We recall that, for a polynomial dynamical system **x**^*′*^ = **f** (**x**), a polynomial *p*(**x**) is called *a Darboux polynomial* if there exists another polynomial *q*(**x**) such that (*p*(**x**))^*′*^ = *q*(**x**)*p*(**x**).

### Lemma B.2

*If none of β, γ, σ is zero, the only irreducible Darboux polynomial of* (B.2) *is X*_0_.

*Proof*. We first note that *X*_0_ is indeed a Darboux polynomial. We will now prove that there are no other irreducible Darboux polynomials for (B.2). Let **X** = (*X*_0_, …, *X*_*n*+1_) and 𝕂 = ℂ (*β, γ, σ*). Assume that *p*(**X**) ∈ 𝕂 [**X**] is a Darboux polynomial of (B.2) with the corresponding eigenpolynomial being *q*(**X**). Since the Lie derivative with respect to (B.2) increases the degree by at most one, we have deg *q*(**X**) ∈ {0, 1}. We consider these cases separately.

Assume that deg *q*(**X**) = 1, so *q*(**X**) = *ℓ*(**X**) + *C*, where *ℓ*(**X**) is a linear form and *C* ∈ 𝕂. Let *p*_0_(**X**) be the homogeneous component of *p*(**X**) of the highest total degree. Then the highest homogeneous component of (*p*(**X**))^*′*^ is 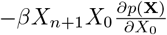. On the other hand, it is equal to *p*_0_(**X**)*ℓ*(**X**), this implies that *ℓ*(**X**) = *AX*_*n*+1_ for some *A* ∈ 𝕂. If *p*(**X**) ≠ *X*_0_, then its irreducibility implies that there exists a monomial not involving *X*_0_. Let us take, among all such monomials, a monomial *m*(**X**) of the highest total degree. Then (*p*(**X**))^*′*^ must involve *X*_*n*+1_*m*(**X**) but this is impossible since all the monomials of degree higher than *m* are divisible by *X*_0_ and taking Lie derivative with respect to (B.2) preserves divisibility by *X*_0_. So, we have arrived to a contradiction for the case deg *q*(**X**) = 1.

Now we consider the case deg *q*(**X**) = 0, so *q* = *C* ∈ 𝕂. Let *i* will be the smallest index such that *X*_*i*_ appears in *p*(**X**). If *i* = 0, consider a monomial *m*(**X**) in *p*(**X**) involving *X*_0_ and of the highest total degree among such monomials. Then *p*^*′*^(**X**) will involve a monomial *X*_*n*+1_*m*(**X**) but this monomial is of higher degree than *m*(**X**), so it cannot appear in *cp*(**X**). If *i >* 0, then (*p*(**X**))^*′*^ must involve *X*_*i*−1_ (since it appears in 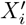) which contradicts the fact that (*p*(**X**))^*′*^ and *p*(**X**) are proportional.

### Corollary

*If none of β, γ, σ is zero, the system* (B.2) *does not have rational first integrals*.

*Proof*. If 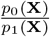 is a rational first integral for *p*_0_, *p*_1_ ∈ 𝕂 [**X**] (in the notation of the proof of Lemma B.2), then the zero set *p*_0_(**X**) −*cp*_1_(**X**) = 0 is an invariant hypersurface for (B.2) for every *c* ∈ 𝕂, so *p*_0_(**X**) −*cp*_1_(**X**) is a Darboux polynomial for every *c* which is impossible since *X*^*m*^ for *m* ∈ ℤ_*>*0_ are the only Darboux polynomials of (B.2).

After we established that the use of input-output equations for all the models in the series Σ_*n*_ is legitimate, we will investigate the structure of the input-output equation of the model.

### Lemma B.3

*The order of the input-output equation of* (B.1) *(and, consequently, of* (B.2)*) is equal to n*+2.

*Proof*. We define polynomials *y*_0_, …, *y*_*n*+2_ ∈ ℂ (*β, γ, σ*)[*S, E*_1_, …, *E*_*n*_, *I*] by *y*_0_ = *σE*_*n*_ and *y*_*i*+1_ is the Lie derivative of *y*_*i*_ with respect to (B.1) for every 0 *< i* ⩽ *n* + 2. Then, for every 0 ⩽ *i* ⩽ *n* + 2, *y*_*i*_ will be the expression of *y*^(*i*)^ in terms of the states and parameters of the model. Since *y*_0_, …, *y*_*n*+2_ are *n* + 3 polynomials in *n* + 2 variables *S, E*_1_, …, *E*_*n*_, *I* over a field ℂ (*β, γ, σ*), they are algebraically dependent over this field. Therefore, the order of the minimal equation satisfied by *y* over ℂ (*β, γ, σ*) does not exceed *n* + 2.

Now we will prove that the order is at least *n* + 2. We will prove this by showing that *y*_0_, …, *y*_*n*+1_ are algebraically independent over ℂ (*β, γ, σ*). We order variables as *S, E*_1_, …, *E*_*n*_, *I* and number them by integers from 0 to *n* + 1. One can show by induction that the highest variable in *y*_*i*_ is the *i*-th variable in the list. Therefore, the Jacobian of these polynomials is upper-triangular with nonzero entries on the diagonal, so they are algebraically independent.

### Lemma B.4

*We will use the notation y*_0_, …, *y*_*n*+2_ *introduced in the beginning of the proof of Lemma B.3. Assume that n* ⩾ 2. *Then there exist linear forms ℓ*_0_, *ℓ*_1_, *ℓ*_2_ ∈ 𝕂 [*y*_0_, …, *y*_*n*−1_] *such that*

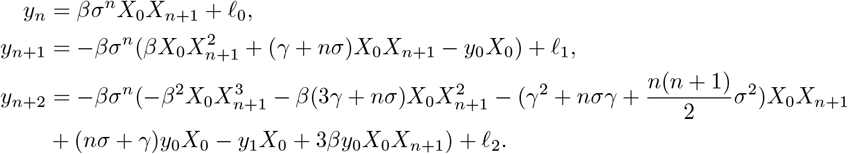

*Proof*. Let *V* be the 𝕂-subspace of 𝕂[*X*_0_, …, *X*_*n*+1_] spanned by linear forms *X*_*i*+1_ − *X*_*i*_ for *i* = 0, …, *n* − 1. We observe that *y* ∈ *V*. Furthermore, if the linear part of *P* ∈ 𝕂[*X*_0_, …, *X*_*n*+1_] belongs to *V*, then the same is true for the linear part of *P*^*′*^ due to the form of the vector field (B.2). Therefore, the 𝕂-space spanned by the linear forms *y*_0_, …, *y*_*n*−1_ belongs to *V*_0_.

In order to establish the reverse inclusion, we will prove the following statement by induction on *k*. For every 0 ⩽ *k < n*, we claim that

- the linear span of *y*_0_, …, *y*_*k*_ contains *X*_*n*−*i*_ − *X*_*n*−*i*−1_ for *i* = 0, …, *k*;
- 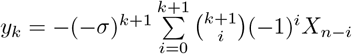

The statement is trivial for *k* = 0. Assume that it has been proven for some *k < n*− 1. In order to prove the first statement, we take *X*_*n*−*k*_ −*X*_*n*−*k*−1_ which belongs to the span of *y*_0_, …, *y*_*k*_ by the induction hypothesis. Then its derivative belongs to the span of *y*_0_, …, *y*_*k*+1_ and we can write

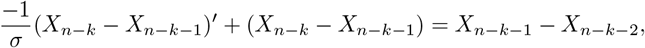

so *X*_*n*−*k*−1_ *X*_*n*−*k*−2_ belongs to the span of *y*_0_, …, *y*_*k*+1_. Now we prove the induction step for the second statement:

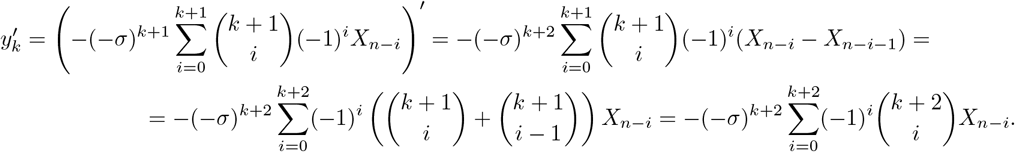

Now we consider

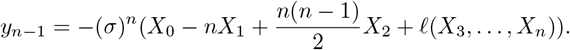

We note that the first three derivatives of the linear form *ℓ*(*X*_3_, …, *X*_*n*_) will be linear forms in *X*_0_, …, *X*_*n*_, so the nonlinear part of *y*_*n*_, *y*_*n*+1_, *y*_*n*+2_ is completely determined by the Lie derivatives of the first three terms in the above formula. The statement of the lemma is thus obtained by computing these Lie derivatives explicitly and then separating the linear part (which will together with the derivative of *ℓ* belong to the span of *y*_0_, …, *y*_*n*−1_). This explicit computation (which we verified with Maple) finishes the proof.

*Proof of Theorem B.1*. First, we consider separately the case *n* = 1. We use `StructuralIdentifiability.jl` to find the input-output equation and verify that its coefficients are indeed globally identifiable (i.e., the Wronskian check). The equation is too long to be displayed here but it contains the following terms:

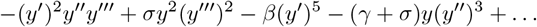

implying that *β, γ, σ* are all globally identifiable.

Consider *ℓ*_0_, *ℓ*_1_, *ℓ*_2_ from Lemma B.4. Consider polynomials *A*_*i*_ := *y*_*n*+*i*_ − *ℓ*_*i*_ ∈ 𝕂[*X*_0_, …, *X*_*n*+1_] for *i* = 0, 1, 2. By Lemma B.4, we have *A*_0_, *A*_1_, *A*_2_ ∈ ℂ(*β, γ, σ*)[*y*_0_, *y*_1_, *X*_0_, *X*_*n*+1_]. We use Gröbner bases to find relations between *A*_0_, *A*_1_, *A*_2_, *y*_0_, *y*_1_ over ℂ(*β, γ, σ*) (corresponding Maple script is available in the supplementary code), and we find that there is a unique irreducible relation, we will denote it *P* (*A*_0_, *A*_1_, *A*_2_, *y*_0_, *y*_1_), and this relation is homogeneous of degree three with respect to *A*_0_, *A*_1_, *A*_2_. Substituting back the expression of *A*’s in terms of *y*’s, we find that

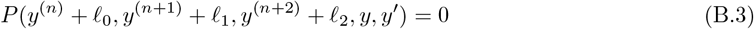

is an input-output equation for (B.1). Lemma B.3 implies that it is of the lowest possible order. Furthermore, since *P* was irreducible and (B.3) is obtained from it by a linear change of variables, this equation is irreducible. Therefore, (B.3) is the minimal irreducible input-output equation for (B.1).

Since *P* (*A*_0_, *A*_1_, *A*_2_, *y*_0_, *y*_1_) is homogeneous of degree three in *A*_0_, *A*_1_, *A*_2_, the homogeneous degree three part with respect to *y*^(*n*)^, *y*^(*n*+1)^, *y*^(*n*+2)^ of (B.3) will be precisely *P* (*y*^(*n*)^, *y*^(*n*+1)^, *y*^(*n*+1)^, *y, y*^*′*^). Here are the relevant monomials from this polynomial:

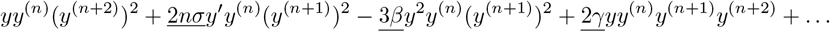

By Corollary B.2.1, the model does not have rational first integrals, so by [40, Theorem 4.7] the underlined coefficients are globally identifiable. Thus, parameters *β, γ, σ* are globally identifiable.

Now we will show that the states are globally identifiable. We will prove that *E*_*n*−*k*_ is identifiable for *k* = 0, …, *n* − 1 by induction on *k*. For the base case *k* = 0, since *y* = *σE*_*n*_ and *σ* is identifiable *E*_*n*_ is identifiable. Assume that we have established the identifiability of *E*_*n*−*k*_ for some *k < n* − 1. Then 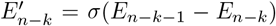 is identifiable as well. Identifiability of *σ* and *E*_*n*−*k*_ implies the identifiability of *E*_*n*−*k*−1_ proving the induction step. Similarly, by considering 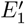, we conclude that *SI* is identifiable. By differentiating one more time, we can conclude the identifiability of

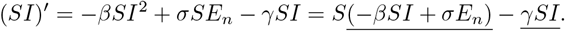

We note that the underlined expressions in the above formula are combinations of identifiable functions and are thus identifiable. Since *βSI* +*σE*_*n*_ is not identically zero for a generic solution of (B.1), we conclude that *S* is observable. Thus, *I* is as observable as well. Finally, knowing *N* implies that *R* is also identifiable.

## Footnotes

1 Theorem 4.2 in the arXiv version

2 Lemma 4.6 in the arXiv version

