## Supplementary material for "Uncovering identifiability of epidemiological models: basic reproduction number and complementary data streams": supp

**S9 Idea Behind Conjecure** 2 **22**

### S1 Proof of Theorem 3.1

*Proof.* We begin by reiterating the SIR model for convenience:

$$\begin{cases} S' = -\beta \dfrac{SI}{N}, \\ I' = \beta \dfrac{SI}{N} - \gamma I, \\ R' = \gamma I, \end{cases} \tag{S1.1}$$

The $R'$ equation is decoupled from the $S'$ and $I'$ equations in, (S1.1) so we begin by analyzing the following reduced system:

$$\begin{cases} S' = -\beta \dfrac{SI}{N} \\ I' = \beta \dfrac{SI}{N} - \gamma I \end{cases} \tag{S1.2}$$

Let $y = \beta \frac{SI}{N}$ be the observation.

#### (i) Identifiability of parameters

The following input-output equation can be obtained by using `StructuralIdentifiability.jl`:

```
N^2*gamma^2*y(t)_0^3*y(t)_2 - N^2*gamma^2*y(t)_0^2*y(t)_1^2
+ N^2*gamma*y(t)_0^2*y(t)_1*y(t)_2 - N^2*gamma*y(t)_0*y(t)_1^3
+ N^2*y(t)_0^2*y(t)_2^2 - 2*N^2*y(t)_0*y(t)_1^2*y(t)_2 + N^2*y(t)_1^4
+ N*beta*gamma^2*y(t)_0^5 + 2*N*beta*gamma*y(t)_0^4*y(t)_1 + 4*N*beta*y(t)_0^4*y(t)_2
- 4*N*beta*y(t)_0^3*y(t)_1^2 + 4*beta^2*y(t)_0^6
```

which can be rewritten as (with $y(t)_0 := y$, $y(t)_1 := y'$, and $y(t)_2 := y''$):

$$\begin{aligned} &N^2\gamma^2 y^3 y'' - N^2\gamma^2 y^2 (y')^2 + N^2\gamma y^2 y' y'' - N^2\gamma y (y')^3 + N^2 y^2 (y'')^2 - 2N^2 y (y')^2 y'' \\ &+ N^2 (y')^4 + N\beta\gamma^2 y^5 + 2N\beta\gamma y^4 y' + 4N\beta y^4 y'' - 4N\beta y^3 (y')^2 + 4\beta^2 y^6 = 0 \end{aligned} \tag{S1.3}$$

Dividing (S1.3) by $N^2\gamma^2$ gives the following monic polynomial:

$$\begin{aligned} &y^3 y'' - y^2 (y')^2 + \frac{1}{\gamma} y^2 y' y'' - \frac{1}{\gamma} y (y')^3 + \frac{1}{\gamma^2} y^2 (y'')^2 - \frac{2}{\gamma^2} y (y')^2 y'' \\ &+ \frac{1}{\gamma^2} (y')^4 + \frac{\beta}{N} y^5 + \frac{2\beta}{N\gamma} y^4 y' + \frac{4\beta}{N\gamma^2} y^4 y'' - \frac{4\beta}{N\gamma^2} y^3 (y')^2 + \frac{4\beta^2}{N^2\gamma^2} y^6 = 0. \end{aligned} \tag{S1.4}$$

We note that, when computing the input-output equation, `StructuralIdentifiability.jl` checks the condition from (Step 2) of Section 2.3 of the main text. More precisely, it checks the nonsingularity of the appropriate Wronskian (see Appendix A and [4, Section 5.3]) by evaluating it at a random point. If established, nonsingularity at a point implies that the Wronskian is nonsingular on an open dense set, thus allowing us to use the coefficients of the input-output equation to generate all the identifiable combinations. This check was successfully passed for all the models considered in this paper, so we will not mention it explicitly in the subsequent proofs.

Suppose another set of parameters $(\beta^\star, \gamma^\star, N^\star)$ can generate the same monic polynomial (S1.4). Then we have:

$$\frac{1}{\gamma} = \frac{1}{\gamma^\star} \quad \text{and} \quad \frac{\beta}{N} = \frac{\beta^\star}{N^\star}$$

from which it follows that $\gamma = \gamma^*$, hence $\gamma$ is globally identifiable. Furthermore, the quotient $\frac{\beta}{N}$ is always identifiable. On the other hand, $\beta$ and $N$ always appear in the coefficients of the IO-equation as a quotient, so they are not identifiable themselves.

The input-output equation can also be obtained using DAISY. In this example, it happens that the input-output equation obtained through DAISY (with $y_1 := y$, $\mathrm{df}(y_1, t) := y'$, and $\mathrm{df}(y_1, t, 2) := y''$) is identical to (S1.3):

```
aa_(1) := df(y1,t,2)**2*y1**2*n**2 - 2*df(y1,t,2)*df(y1,t)**2*y1*n**2
+ df(y1,t,2)*df(y1,t)*y1**2*gamma*n**2 + 4*df(y1,t,2)*y1**4*beta*n
+ df(y1,t,2)*y1**3*gamma**2*n**2 + df(y1,t)**4*n**2
 - df(y1,t)**3*y1*gamma*n**2 - 4*df(y1,t)**2*y1**3*beta*n
 - df(y1,t)**2*y1**2*gamma**2*n**2 + 2*df(y1,t)*y1**4*beta*gamma*n + 4*y1**6*beta**2
+ y1**5*beta*gamma**2*n$
```

#### (ii) Identifiability of state variables

We reparameterize model (S1.2) by setting $a = \frac{\beta}{N}$. The resulting model is given by:

$$\begin{cases} S' = -aSI \\ I' = aSI - \gamma I \end{cases} \tag{S1.5}$$

Since $y = aSI$ is the observation, it follows from the first equation in (S1.5) that $S'$ is identifiable. Since $S'$ is identifiable, this implies that $S + c$ for some constant is also identifiable. Hence, to show $S$ is identifiable, it is sufficient to show that $c$ is identifiable. We now consider the model (S1.2) with two observations, namely: $y_1 = \beta \frac{SI}{N}$ and $y_2 = S + c$. The system of input-output equation obtained using `StructuralIdentifiability.jl` is:

```
N*c*gamma*y1(t)_0 + N*c*y1(t)_1 - N*gamma*y1(t)_0*y2(t)_0 - N*y1(t)_0^2
- N*y2(t)_0*y1(t)_1 + beta*c^2*y1(t)_0 - 2*beta*c*y1(t)_0*y2(t)_0
+ beta*y1(t)_0*y2(t)_0^2
y1(t)_0 + y2(t)_1
```

which can be rewritten as:

$$\begin{aligned} Nc\gamma y_1 + Ncy_1' - N\gamma y_1 y_2 - Ny_1^2 - Ny_2 y_1' + \beta c^2 y_1 - 2\beta c y_1 y_2 + \beta y_1 y_2^2 = 0 \\ y_1 + y_2' = 0. \end{aligned} \tag{S1.6}$$

We divide the first equation in (S1.6) by $N$ to get:

$$N\gamma y_1 + cy_1' - \gamma y_1 y_2 - y_1^2 - y_2 y_1' + \frac{\beta}{N} c^2 y_1 - \frac{2\beta c}{N} y_1 y_2 + \frac{\beta}{N} y_1 y_2^2 = 0, \tag{S1.7}$$

from which it follows that $c$ is globally identifiable and thus $S$ is also globally identifiable.

Since $a = \frac{\beta}{N}$ and $S$ are identifiable and $y = aSI$ is the observation, it must be that $I$ is also identifiable. However, since $N$ is unknown and $R = N - S - I$, $R$ remains nonidentifiable. Hence, $S$ and $I$ are globally identifiable; however $R$ is nonidentifiable.

#### (iii) Identifiability with one additional complementary data point

Since the SIR model (S1.2) is a closed system with $N = S(t) + I(t) + R(t)$ constant for all time $t$, and since $S(t)$ and $I(t)$ are identifiable, knowing $R(t)$ at any single point in time identifies $N$, which in turn makes $\beta$ globally identifiable. Moreover, with $N$ known and $S$ and $I$ globally identifiable, $R = N - S - I$ is also globally identifiable. Hence, the SIR model (S1.2) becomes globally identifiable. □

#### S2 Idea Behind Conjecture 1

Since $R'$ equation is decoupled from the rest of the equations in (3.3), we rewrite the model as:

$$\begin{cases} S' = -\beta S \sum_{j=1}^{n} I_j, \\ I_1' = \beta S \sum_{j=1}^{n} I_j - \zeta I_1, \\ I_i' = \zeta I_{i-1} - \zeta I_i, \quad i = 2, \dots, n-1 \\ I_n' = \zeta I_{n-1} - \zeta I_n, \end{cases} \tag{S2.1}$$

***n* = 1**

For $n = 1$, model (S2.1) reduces to the SIR model (3.1). The proof for this scenario is given in Section S1. For $y = \beta S \sum_{j=1}^{n} I_j$, we check the identifiability for $n = 2, ...5$ using `StructuralIdentifiability.jl`.

***n* = 2**

```
ode = @ODEmodel(
    S'(t) = -beta*(I1(t)+I2(t))*S(t),
    I1'(t) = beta*(I1(t)+I2(t))*S(t) - zeta*I1(t),
    I2'(t) = zeta*I1(t) - zeta*I2(t),
    y(t) = beta*(I1(t)+I2(t))*S(t)
)
```

`assess_identifiability(ode)` gives:

```
S(t)  => :globally
I1(t) => :globally
I2(t) => :globally
beta  => :globally
zeta  => :globally
```

***n* = 3**

```
ode = @ODEmodel(
    S'(t) = -beta*(I1(t)+I2(t)+I3(t))*S(t),
    I1'(t) = beta*(I1(t)+I2(t)+I3(t))*S(t) - zeta*I1(t),
    I2'(t) = zeta*I1(t) - zeta*I2(t),
    I3'(t) = zeta*I2(t) - zeta*I3(t),
    y(t) = beta*(I1(t)+I2(t)+I3(t))*S(t)
)
```

`assess_identifiability(ode)` gives:

```
S(t)  => :globally
I1(t) => :globally
I2(t) => :globally
I3(t) => :globally
beta  => :globally
zeta  => :globally
```

$\boldsymbol{n = 4}$

```
ode = @ODEmodel(
    S'(t) = -beta*(I1(t)+I2(t)+I3(t)+I4(t))*S(t),
    I1'(t) = beta*(I1(t)+I2(t)+I3(t)+I4(t))*S(t) - zeta*I1(t),
    I2'(t) = zeta*I1(t) - zeta*I2(t),
    I3'(t) = zeta*I2(t) - zeta*I3(t),
    I4'(t) = zeta*I3(t) - zeta*I4(t),
    y(t) = beta*(I1(t)+I2(t)+I3(t)+I4(t))*S(t)
)
```

`assess_identifiability(ode)` gives:

```
 S(t)  => :globally
  I1(t) => :globally
  I2(t) => :globally
  I3(t) => :globally
  I4(t) => :globally
  beta  => :globally
  zeta  => :globally
```

$\boldsymbol{n = 5}$

```
ode = @ODEmodel(
ode = @ODEmodel(
    S'(t) = -beta*(I1(t)+I2(t)+I3(t)+I4(t)+I5(t))*S(t),
    I1'(t) = beta*(I1(t)+I2(t)+I3(t)+I4(t)+I5(t))*S(t) - zeta*I1(t),
    I2'(t) = zeta*I1(t) - zeta*I2(t),
    I3'(t) = zeta*I2(t) - zeta*I3(t),
    I4'(t) = zeta*I3(t) - zeta*I4(t),
    I5'(t) = zeta*I4(t) - zeta*I5(t),
    y(t) = beta*(I1(t)+I2(t)+I3(t)+I4(t)+I5(t))*S(t)
)
```

`assess_identifiability(ode)` gives:

```
  S(t)  => :globally
  I1(t) => :globally
  I2(t) => :globally
  I3(t) => :globally
  I4(t) => :globally
  I5(t) => :globally
  beta  => :globally
  zeta  => :globally
```

In each of these scenarios, the total population $N$ is assumed to be known. Since $R = N - S - \sum_{j=1}^{n} I_j$ can be expressed in terms of globally identifiable quantities, it is also globally identifiable. Thus, the model is globally identifiable for $n = 1, 2, 3, 4, 5$ (with high probability).

#### S3 Proof of Theorem 3.3

*Proof.* We begin by reiterating the SEIRD model (3.4):

$$\begin{cases} S' = -\beta \dfrac{SI}{S+I+R} \\ I' = \beta \dfrac{SI}{S+I+R} - \widetilde{\gamma} I \\ R' = (1-\phi_d)\widetilde{\gamma} I \\ D' = \phi_d \widetilde{\gamma} I, \end{cases} \tag{S3.1}$$

Since we assume that the total initial population $N(0)$ is known, without loss of generality, let $N = 1$. Hence, $S + I + R$ can be rewritten as $N - D$. Thus, we decoupled the $R'$ equation from the rest of the equation in (S3.1) to get the following model:

$$\begin{cases} S' = -\beta \dfrac{SI}{1-D} \\ I' = \beta \dfrac{SI}{1-D} - \widetilde{\gamma} I \\ D' = \phi_d \widetilde{\gamma} I, \end{cases} \tag{S3.2}$$

Let new mortality be the observation with $y = \phi_d \widetilde{\gamma} I$.

##### (i) Identifiability of parameters

Since $N$ is assumed to be known, without loss of generality, let $N = 1$. Then, the following input-output equation can be obtained from `StructuralIdentifiability.jl` (for simplicity `gamma` and `phi` are used instead of $\widetilde{\gamma}$ and $\phi_d$, respectively):

```
beta*gamma*y(t)_0^3*y(t)_3 - 4*beta*gamma*y(t)_0^2*y(t)_1*y(t)_2
+ 3*beta*gamma*y(t)_0*y(t)_1^3 + beta*y(t)_0^2*y(t)_1*y(t)_3 - beta*y(t)_0^2*y(t)_2^2
- 2*beta*y(t)_0*y(t)_1^2*y(t)_2  + 2*beta*y(t)_1^4 - gamma^2*phi*y(t)_0^3*y(t)_3
+ 4*gamma^2*phi*y(t)_0^2*y(t)_1*y(t)_2
- 3*gamma^2*phi*y(t)_0*y(t)_1^3 - gamma*phi*y(t)_0^2*y(t)_1*y(t)_3
+ 2*gamma*phi*y(t)_0^2*y(t)_2^2 - gamma*phi*y(t)_1^4
```

which can be written as

$$\begin{aligned} &\beta\widetilde{\gamma} y^3 y''' - 4\beta\widetilde{\gamma} y^2 y' y'' + 3\beta\widetilde{\gamma} y (y')^3 + \beta y^2 y' y''' - \beta y^2 (y'')^2 - 2\beta y (y')^2 y'' + 2\beta (y')^4 - \widetilde{\gamma}^2 \phi_d y^3 y''' \\ &+ 4\widetilde{\gamma}^2 \phi_d y^2 y' y'' - 3\widetilde{\gamma}^2 \phi_d y (y')^3 - \widetilde{\gamma}\phi_d y^2 y' y''' + 2\widetilde{\gamma}\phi_d y^2 (y'')^2 - \widetilde{\gamma}\phi_d (y')^4 = 0 \end{aligned} \tag{S3.3}$$

The monic polynomial corresponding to the input-output equation (obtained by dividing by $\beta\widetilde{\gamma} - \widetilde{\gamma}^2\phi$, which is equivalent to $\widetilde{\gamma}(\beta - \widetilde{\gamma}\phi)$) and grouping like terms in the above output) is:

$$y^3 y''' - \frac{1}{\widetilde{\gamma}} y^2 y' y''' + 3y(y')^3 - \frac{1}{\widetilde{\gamma}} y^2 (y'')^2 - \frac{2\beta}{\beta\widetilde{\gamma} - \widetilde{\gamma}^2\phi_d} y (y')^2 y'' + \frac{2\beta - \widetilde{\gamma}\phi_d}{\beta\widetilde{\gamma} - \widetilde{\gamma}^2\phi_d} (y')^4 - 4y^2 y' y'' = 0 \tag{S3.4}$$

Suppose another set of parameters $(\beta^*, \widetilde{\gamma}^*, \phi^*)$ can generate the same monic polynomial (S3.4). Then we have:

$$\frac{1}{\widetilde{\gamma}} = \frac{1}{\widetilde{\gamma}^*}, \quad \frac{2\beta}{\beta\widetilde{\gamma} - \widetilde{\gamma}^2\phi_d} = \frac{2\beta^*}{\beta^*\widetilde{\gamma}^* - (\widetilde{\gamma}^2)^*\phi_d^*} \quad \text{and} \quad \frac{2\beta - \widetilde{\gamma}\phi_d}{\beta\widetilde{\gamma} - \widetilde{\gamma}^2\phi_d} = \frac{2\beta^* - \widetilde{\gamma}^*\phi_d^*}{\beta^*\widetilde{\gamma}^* - (\widetilde{\gamma}^2)^*\phi_d^*},$$

from which it follows that

$$\widetilde{\gamma} = \widetilde{\gamma}^* \quad \text{and} \quad \frac{\beta}{\phi_d} = \frac{\beta^*}{\phi_d^*}.$$

hence $\widetilde{\gamma}$ and $\frac{\beta}{\phi_d}$ are globally identifiable. Furthermore, the above formula shows that $\beta$ and $\phi_d$ are nonidentifiable since any scaling transformation $\beta^* = \lambda\beta, \phi_d^* = \lambda\phi_d$ with nonzero $\lambda$ yields another parameter value giving the same IO-equation in an arbitrarily small neighborhood of the original parameter values.

We note that the input-output equation obtained from DAISY (given below) is equivalent to the one given in (S3.3).

```
df(y1,t,3)*df(y1,t)*y1**2*( - beta + gamma*phi)
+ df(y1,t,3)*y1**3*gamma*( - beta + gamma*phi)
+ df(y1,t,2)**2*y1**2*(beta - 2*gamma*phi) + 2*df(y1,t,2)*df(y1,t)**2*y1*beta
+ 4*df(y1,t,2)*df(y1,t)*y1**2*gamma*(beta - gamma*phi)
+ df(y1,t)**4*( - 2*beta + gamma*phi) + 3*df(y1,t)**3*y1*gamma*( - beta + gamma*phi)
```

#### (ii) Identifiability of state variables

Since $D'$ is the observation, we choose known $M$ with the following property: $M'(t) = D'(t)$. Hence $M(t) = D(t) + c$, where c is some constant. Thus, model (S3.2) can be rewritten as:

$$\begin{cases} S' = -\beta\dfrac{SI}{1-D-c} \\ I' = \beta\dfrac{SI}{1-D-c} - \widetilde{\gamma}I \\ M' = \phi_d\widetilde{\gamma}I, \end{cases} \tag{S3.5}$$

We reparameterize model (S3.5) through the variable transformations $(X_1, X_2, X_3) := (M, \phi_d S, \phi_d I)$ and the parameter transformation $a = \beta/\phi_d$, while $\widetilde{\gamma}$ and $c$ remain unchanged. The resulting model is as follows:

$$\begin{cases} X_1' = \widetilde{\gamma}X_3, \\ X_2' = -\dfrac{aX_2(t)X_3}{1-X_1-c}, \\ X_3' = \dfrac{\widetilde{\gamma}X_1X_3 + aX_2(t)X_3(t) + \widetilde{\gamma}cX_3 - \widetilde{\gamma}X_3(t)}{1-X_1-c}, \end{cases} \tag{S3.6}$$

Recall that $\widetilde{\gamma}$ and $c$ were structurally identifiable. Since $M(t)$ is known, $X_1(t)$, which equals $M(t)$, is known, and so is its derivative $X_1'$. Since $X_1'$ and $\widetilde{\gamma}$ are known in the first equation of (S3.6), it follows that $X_3$ is known. Since $X_3$ is known, this implies that $X_3'$ is known. It follows from the third equation of (S3.6) that $X_2$ is known. Since $X_2 = \phi_d S$ and $X_3 = \phi_d I$, this implies that the quantities $\phi_d S(t)$ and $\phi_d I(t)$ of the model (S3.1) are structurally identifiable with the observation of new mortality ($\phi_d\widetilde{\gamma}I$).

Finally, we note that $I(t)$, $S(t)$, and $R(t)$ are not themselves nonidentifiable. Indeed, for any nonzero $\lambda$, the transformation (noting that the $R$ transformation follows from $R = N - S - I$ being conserved)

$$S \to \lambda S, \;\; I \to \lambda I, \;\; R \to R + (1-\lambda)(S+I), \;\; \beta \to \frac{\beta}{\lambda}, \;\; \phi_d \to \frac{\phi_d}{\lambda}$$

leaves the output of the model, $\phi_d \tilde{\gamma} I$ invariant while allowing us to obtain infinitely many different trajectories for $S$, $I$, and $R$ with the initial conditions and parameter values in an arbitrarily small neighborhood of the original values.

#### (iii) Identifiability with one additional complementary data point

Since $\phi_d$ is a constant, and both $\phi_d S(t)$ and $\phi_d I(t)$ are identifiable, it implies that having any single data point in $S(t)$ or $I(t)$ curve (in addition to new mortality), makes the parameter $\phi_d$ identifiable, which then makes $\beta$ identifiable. Moreover, with $\phi_d$ identifiable, the identifiability of $\phi_d S(t)$ and $\phi_d I(t)$ implies that $S(t)$ and $I(t)$ are also globally identifiable.

Furthermore, since $N(t) = S(t) + I(t) + R(t) - D(t)$, we have:

$$S(t) + I(t) = N(t) + D(t) - R(t).$$

Since $N(t = 0)$ is known and mortality is the only mechanism for population depletion, $N(t) + D(t) = N(0)$ is known. Therefore, if $R(t_i)$ is known at time $t_i$, then

$$S(t_i) + I(t_i) = N(0) - R(t_i)$$

is also known since the right hand side is a known quantity. Moreover, because $\phi_d S(t)$ and $\phi_d I(t)$ are identifiable, so is $\phi_d S(t_i) + \phi_d I(t_i) = \phi_d [S(t_i) + I(t_i)]$. Since the product of two factors, $\phi_d(S(t_i) + I(t_i))$, is identifiable and one factor of the product, $(S(t_i) + I(t_i))$, is known, the other factor, $\phi_d$, must also be identifiable. This shows that in addition to new mortality, having even a single noise-free data point in the $R(t)$ curve also makes the model (S3.1) structurally identifiable. □

#### S4 Proof of Theorem 3.4

*Proof.* We rewrite the cholera model (3.6) for convenience:

$$\begin{cases} S' = -\beta_W SW - \beta_I SI, \\ I' = \beta_W SW + \beta_I SI - \gamma I, \\ W' = \alpha I - \zeta W, \\ R' = \gamma I. \end{cases} \tag{S4.1}$$

Notice that the $R'$ equation is decoupled from the rest of the equations in (S5.1), hence we rewrite the model as:

$$\begin{cases} S' = -\beta_W SW - \beta_I SI, \\ I' = \beta_W SW + \beta_I SI - \gamma I, \\ W' = \alpha I - \zeta W. \end{cases} \tag{S4.2}$$

Let $y = W$ be the observation.

##### (i) Identifiability of parameters

We use `StructuralIdentifiability.jl` to obtain the input-output equation for the model. For convenience, we do not display it as a single equation, but give the list of the monomials with respect to $y$ and its derivatives indicating the corresponding coefficients (after normalization):

1. $(y'')^2$ with coefficient: 1;
2. $y'y'''$ with , coefficient $-1$;
3. $(y')^2 y''$ with coefficient $-\frac{\beta_I}{\alpha}$;
4. $y'y'$ with coefficient $\frac{\alpha\beta_W + \beta_I\zeta}{\beta_I}$;
5. $(y')^3$ with coefficient $\frac{-\beta_I\gamma - \beta_I\zeta}{\alpha}$;
6. $yy'''$ with coefficient $\frac{-\alpha\beta_W - \beta_I\zeta}{\beta_I}$;
7. $yy'y''$ with coefficient $-2\frac{\alpha\beta_W + \beta_I\zeta}{\alpha}$;
8. $(y')^2$ with coefficient $\frac{\alpha\beta_W\gamma + \alpha\beta_W\zeta + \beta_I\zeta^2}{\beta_I}$;
9. $yy''$ with coefficient $\frac{-\alpha\beta_W\gamma - \alpha\beta_W\zeta - \beta_I\zeta^2}{\beta_I}$;
10. $y^2 y''$ with coefficient $\frac{-\alpha^2\beta_W^2 - 2\alpha\beta_I\beta_W\zeta - \beta_I^2\zeta^2}{\alpha\beta_I}$;
11. $y^3$ with coefficient $\frac{-\alpha^2\beta_W^2\gamma\zeta - 2\alpha\beta_I\beta_W\gamma\zeta^2 - \beta_I^2\gamma\zeta^3}{\alpha\beta_I}$;
12. $y(y')^2$ with coefficient $\frac{-2\alpha\beta_W\gamma - 2\alpha\beta_W\zeta - 3\beta_I\gamma\zeta - 2\beta_I\zeta^2}{\alpha}$;
13. $y^2 y'$ with coefficient $\frac{-\alpha^2\beta_W^2\gamma - \alpha^2\beta_W^2\zeta - 4\alpha\beta_I\beta_W\gamma\zeta - 2\alpha\beta_I\beta_W\zeta^2 - 3\beta_I^2\gamma\zeta^2 - \beta_I^2\zeta^3}{\alpha\beta_I}$.

As was explained in the previous proofs, the software checks the Wronskian condition, so these coefficients give a complete set of identifiable combinations. These expressions are already relatively large and not very convenient to work with, but we can ask software to propose a simpler complete set of identifiable combinations [3] which will be:

$$\gamma + \zeta, \quad \gamma\zeta, \quad \frac{\alpha}{\beta_I}, \quad \frac{\alpha\beta_W + \beta_I\zeta}{\beta_I} = \zeta + \frac{\alpha}{\beta_I}\beta_W. \tag{S4.3}$$

The algorithm behind this computation is randomized, so this set is correct with high probability (and, as we will see, it is actually correct), but this is not a complete mathematical proof so far. To give a complete argument, we will use software [2] (implementing [7, Algorithm 1.10]) to propose expressions of (S4.3) in terms of the coefficients of the IO-equation. Denoting the $i$-th coefficient of the IO-equation from the list above by $c_i$, we obtain

$$\gamma + \zeta = \frac{c_5}{c_3}, \qquad \frac{\alpha}{\beta_I} = \frac{-1}{c_3},$$

$$\gamma\zeta = \frac{-c_3c_8 + c_4c_5}{c_3}, \qquad \zeta + \frac{\alpha}{\beta_I}\beta_W = c_4.$$

While the algorithm used in this computation is also randomized, the result can be checked by direct substitution. This calculation shows that (S4.3) are indeed identifiable. In order to show that they give a complete set (that is, any other identifiable combination can be obtained from them using arithmetic operations), it is sufficient to express the coefficients of the IO-equation in terms of (S4.3). We do this again using [2] and verifying the result by direct substitution. We will omit the details of the calculations here, they are available in the supplementary source code.

Using the obtained identifiable combinations, we can now prove the claimed statements about identifiability of parameters. Non-identifiability of $\alpha$ and $\beta_I$ follows from the fact that, for any $\lambda \in \mathbb{R}$, the transformation

$$\alpha \to \lambda\alpha \quad \text{and} \quad \beta_I \to \beta_I$$

does not change the values of (S4.3). Thus, it yields infinitely many possible values of $\alpha$ and $\beta$ giving the same values of identifiable combinations.

In order to establish local identifiability of $\gamma, \zeta$, and $\beta_W$, we observe that $\gamma$ and $\zeta$ can be found, up to a swap, as the roots of an equation $X^2 - (\gamma + \zeta)X + \gamma\zeta = 0$ with identifiable coefficients. Therefore, they are locally identifiable. Based on this, we can show that $\beta_W$ is locally identifiable as it can be expressed in terms of locally and globally identifiable quantities:

$$\beta_W = \frac{(\zeta + \frac{\alpha}{\beta_I}\beta_W) - \zeta}{\alpha/\beta_I}.$$

Finally, we will show that $\gamma, \zeta$, and $\beta_W$ are not globally identifiable. A direct computation shows that the following transformation

$$\alpha \to \alpha,\ \beta_I \to \beta_I,\ \beta_W \to \beta_W + \frac{\zeta - \gamma}{\alpha/\beta_I},\ \gamma \to \zeta,\ \zeta \to \gamma$$

does not change the values of (S4.3) but it changes the values of $\gamma, \zeta, \beta_W$, thus yielding a different set of parameter values giving the same values of the identifiable combinations.

#### (ii) Identifiability of state variables

We reparameterize model (S5.2) through the following parameter and state variable and transformations:

$$(a_1, a_2, a_3) = (\gamma + \zeta, \gamma\zeta, \alpha/\beta_I) \quad \text{and} \quad (X_1, X_2, X_3) = (W, \beta_I I + \beta_W W, \beta_I S),$$

to obtained the following reparameterized model:

$$\begin{cases} X_1' = a_3X_2 - a_4X_1, \\ X_2' = \dfrac{a_1a_4X_1 - a_2X_1 - a_4^2X_1 + a_3X_2X_3 - a_1a_3X_2 + a_3a_4X_2}{a_3}, \\ X_3' = -X_2X_3. \end{cases} \tag{S4.4}$$

First, notice that all parameters of this new model ($a_1, a_2, a_3$, and $a_4$) are identifiable. Since $W$ is the observation and $X_1 = W$, $X_1$ is known, which implies $X_1'$ is known. From the first equation of (S4.4), it follows that $X_2$ is globally identifiable, which implies that $X_2$ is known. From the second equation of (S4.4) it follows that $X_3$ is identifiable as well. Thus, as claimed in the theorem, $\beta_I I + \beta_W W$ and $\beta_I S$ are identifiable.

It now remains to show that $S$, $I$, and $R$ are nonidentifiable. As before (see, e.g., Section S3(ii)), we can construct a one-parameter family (parametrized by $\lambda \in \mathbb{R}$) of output-preserving transformations

$$\alpha \to \lambda\alpha, \;\; \beta_I \to \lambda\beta_I, \;\; S \to \frac{S}{\lambda}, \;\; I \to \frac{I}{\lambda}, \;\; R \to R + (1 - \frac{1}{\lambda})(S + I).$$

This transformation allows us to construct a trajectory with the same output but different initial conditions for $S, I, R$ in an arbitrarily small neighborhood of the original values.

#### (iii) Identifiability with two additional complementary data points

Since $\beta_I S$ is identifiable, if even a single data point in $S(t)$ is known, then $\beta_I$ becomes identifiable. Moreover, since $\beta_I I + \beta_w W$ is identifiable, if a single data point in $I(t)$ curve is also known, then $\beta_w$ is also identifiable. Since $\alpha/\beta_I$ is identifiable, with $\beta_I$ identifiable, so is $\alpha$. Furthermore, because $(\alpha\beta_W + \beta_I\zeta)/\beta_I$ is identifiable, $\zeta$ now becomes identifiable. Finally, since $\zeta\gamma$ is identifiable, this implies that $\gamma$ is also identifiable.

Hence, with $W(t)$ as an observation and a single data point in $S(t)$ and $I(t)$ known, then the model (S4.4) is structurally identifiable. □

### S5 Proof of Theorem 3.5

*Proof.* We rewrite the cholera model (3.6) for convenience:

$$\begin{cases} S' = -\beta_W SW - \beta_I SI, \\ I' = \beta_W SW + \beta_I SI - \gamma I, \\ W' = \alpha I - \zeta W, \\ R' = \gamma I. \end{cases} \tag{S5.1}$$

Notice that the $R'$ equation is decoupled from the rest of the equations in (S5.1), hence we rewrite the model as:

$$\begin{cases} S' = -\beta_W SW - \beta_I SI, \\ I' = \beta_W SW + \beta_I SI - \gamma I, \\ W' = \alpha I - \zeta W. \end{cases} \tag{S5.2}$$

Let $y = \alpha I$ be the observation.

#### (i) Identifiability of parameters

The input-output equation output from `StructuralIdentifiability.jl` is too long and we will not display it here. It contains 52 monomials with respect to $y$ and its derivatives (available in the supplementary code together with the corresponding coefficients).

Using the approach presented in Section S4(i), we establish that the following simple functions can be expressed in terms of these coefficients $\beta_W, \gamma, \zeta, \frac{\alpha}{\beta_I}$ and, vice versa, all the coefficients can be expressed in terms of these functions. This proves the global identifiability of these quantities. The fact that $\alpha$ and $\beta_I$ are not even locally identifiable can be established in the same way as in Section S4(i) by using the transformation $\alpha \to \lambda\alpha,\ \beta_I \to \lambda\beta_I$.

#### (ii) Identifiability of state variables

We reparameterize model (S5.2) through the following parameter and state variable transformations:

$$a = \alpha/\beta_I \quad \text{and} \quad (X_1, X_2, X_3) = (W, \beta_I I, \beta_I S),$$

to obtained the following reparameterized model:

$$\begin{cases} X_1' = aX_2 - \zeta X_1, \\ X_2' = \beta_W X_1 X_3 + X_2 X_3 - \gamma X_2, \\ X_3' = -\beta_W X_1 X_3 - X_2 X_3. \end{cases} \tag{S5.3}$$

Notice that the observation, under the reparameterized model (S5.3), is $a_4 X2(t)$. Since $a_4$ is identifiable, and $a_4 X_2$ is the observation, this implies that $X_2$ is identifiable, which implies that $X_2'$ is also identifiable. Since $\gamma X_2$ is identifiable and $X_2'$ is identifiable, from the second equation of (S5.3), we have that the following term is also identifiable:

$$\beta_W X_1 X_3 + X_2 X_3.$$

Then, since the right-hand side of the third equation of (S5.3) is identifiable, it follows that $X_3'$ is also identifiable, which implies $X_3 + c$ is identifiable. If $c$ is identifiable, then so is $X_3$, and it follows from the third equation of (S5.3) that $X_1$ is identifiable. To prove that $c$ is identifiable, we proceed as follows. Consider the (S5.2) model with observations (a) $\alpha I$ like before but also (b) $X_3 + c = \beta_I S$. Analyzing this input-output equation, using the methodology presented in Section S4(i), shows that $c$ is identifiable. Finally, we can show that $I$, $S$, and $R$ are nonidentifiable similarly to Section S4(ii) by using the following one-parametric family of output-preserving transformations:

$$\alpha \to \frac{\alpha}{\lambda}, \;\; \beta_I \to \frac{\beta_I}{\lambda}, \;\; S \to \lambda S, \;\; I \to \lambda I, \;\; R \to R + (1-\lambda)(I+S).$$

#### (iii) Identifiability with one additional complementary data point

Since $X_1 = \beta_I I$ and $X_2 = \beta_I S$, and $R = N - (S+I)$, it follows that having a single data point in either $S$, $I$, or $R$, allows $\beta_I$ to be identifiable. Since $a = \alpha/\beta_I$ is identifiable, it follows that, with $\beta_I$ identifiable, $\alpha$ is identifiable as well.

Hence, under the observation of $\alpha I$, an addition of one data point in either $S(t)$, $I(t)$, or $R(t)$ is needed for the model (S5.1) to be structurally identifiable. □

### S6 Proof of Theorem 3.6

*Proof.* We rewrite model (3.7) below:

$$\begin{cases} S_1' = -\beta(I_1+I_2)S_1, \\ S_2' = -\beta(I_1+I_2)S_2, \\ I_1' = \beta(I_1+I_2)S_1 - \gamma I_1, \\ I_2' = \beta(I_1+I_2)S_2 - \gamma I_2, \\ R_1' = \gamma I_1, \\ R_2' = \gamma I_2, \end{cases} \tag{S6.1}$$

Since the rest of the model is independent of $R_1$ and $R_2$, we rewrite model (S6.1) as follows:

$$\begin{cases} S_1' = -\beta(I_1+I_2)S_1, \\ S_2' = -\beta(I_1+I_2)S_2, \\ I_1' = \beta(I_1+I_2)S_1 - \gamma I_1, \\ I_2' = \beta(I_1+I_2)S_2 - \gamma I_2, \end{cases} \tag{S6.2}$$

Let $y = I_1 + I_2$ be the observation.

#### (i) Identifiability of parameters

The following input-output equation can be obtained by using `StructuralIdentifiability.jl`:

```
-beta*gamma*y(t)_0^3 - beta*y(t)_0^2*y(t)_1 - y(t)_0*y(t)_2 + y(t)_1^2
```

which can be rewritten as

$$-\beta\gamma y^3 - \beta y^2 y' - yy'' + (y')^2 = 0. \tag{S6.3}$$

Suppose another set of parameters $(\beta^\star, \gamma^\star)$ can produce the same input-output equation. Then, comparing like terms it follows that:

$$\beta = \beta^\star \quad \text{and} \quad \beta\gamma = \beta^\star\gamma^\star.$$

Since $\beta = \beta^\star$, we have that $\beta$ is globally identifiable. With $\beta$ globally identifiable, it follows from $\beta\gamma = \beta^\star\gamma^\star$, that $\gamma$ is also globally identifiable.

We remark that the input-output equation obtained from DAISY (given below) is equivalent to the one given in (S6.3).

```
df(y1,t,2)*y1 - df(y1,t)**2 + df(y1,t)*y1**2*beta + y1**3*beta*gamma
```

#### (ii) Identifiability of state variables

We reparameterize model (S6.2) using $(S, I, R) = (S_1 + S_2, I_1 + I_2, R_1 + R_2)$ to obtain a mass incidence SIR model:

$$\begin{cases} S' = -\beta S I, \\ I' = \beta S I - \gamma I \\ R' = \gamma I, \end{cases} \tag{S6.4}$$

This problem now reduces to the one presented in Appendix A with $N = 1$. Hence, from Appendix A, it follows that $S$ is identifiable, which implies $S_1 + S_2$ is identifiable. Moreover, since $N(0)$ is known and the population is closed, $N(t) = N(0)$ is also known. Furthermore, $R(t) = N(t) - S(t) - I(t)$ is also identifiable, which then implies that $R_1 + R_2$ is also globally identifiable. □

### S7 Proof of Theorem 3.7

We begin by rewriting the model (3.8) for convenience:

$$\begin{cases} I_h' = ab\dfrac{I_v}{H}(H - I_h) - \gamma I_h, \\ I_v' = ac\dfrac{I_h}{H}(V - I_v) - \mu I_v, \end{cases} \tag{S7.1}$$

Let $y = ab\dfrac{I_v}{H}(H - I_h)$ be the observation.

#### (i) Identifiability of parameters

The input-output equation output from `StructuralIdentifiability.jl` is too long, and we will not display it here. We could use the approach presented in Section S4(i) on this long input-output equation to establish which parameters and parameter combinations are globally identifiable. However, we reparameterize the model and augment it with additional identifiable observations. It turns out that the input-output equation of this augmented model is small enough and can be easily manipulated to demonstrate identifiability results algebraically without use of software.

We reparameterize model (S5.2) through the following parameter and state variable transformations:

$$(a_1, a_2) = (ac, Vab) \quad \text{and} \quad X = \frac{I_v}{V},$$

to obtain the following reparameterized model:

$$\begin{cases} I_h' = \dfrac{(-a_2 X I_h + a_2 H X - \gamma H I_h)}{H}, \\ X' = \dfrac{(-a_1 X I_h - \mu H X + a_1 I_h)}{H}. \end{cases} \tag{S7.2}$$

The observation $y$ is now given by:

$$y = \frac{-a_2 X I_h + a_2 H X}{H}. \tag{S7.3}$$

From the first equation of (S7.2) and the observation $y$ given in (S7.3), we have

$$I_h' + \gamma I_h = y. \tag{S7.4}$$

This first-order linear ODE has the solution

$$I_h(t) = e^{-\gamma t}\int_0^t e^{\gamma s} y(s)\, ds + \ C e^{-\gamma t}, \tag{S7.5}$$

where $C = I_h(0)$ is the unknown initial condition. If $\gamma$ is identifiable, then the first term on the right-hand side of (S7.5) is identifiable, which implies $I_h$ would be identifiable if $C$ is identifiable. To verify identifiability of $C$, we augment the system (S7.2) with an auxiliary state $x' = -\gamma x$ representing the exponential term, and introduce the additional observations $y_2 = x$ and $y_3 = I_h - Cx$. Analyzing the input-output system of the following augmented model will not only establish the identifiability of parameters and parameter combinations from the original system, but will also confirm that $C$ is globally identifiable.

```
ode = @ODEmodel(
    Ih'(t) = a*b*Iv(t)/H*(H-Ih(t)) - gamma*Ih(t),
    Iv'(t) = a*c*Ih(t)/H*(V-Iv(t)) - mu*Iv(t),
    x'(t) = -gamma*x(t),
    y1(t) = a*b*Iv(t)/H*(H-Ih(t)),
    y2(t) = x(t),
    y3(t) = Ih(t) - C*x(t)
)
```

The input-output expression obtained from `StructuralIdentifiability.jl` on augumented equation is:

```
C^3*V*a^2*b*c*y2(t)_0^3 - 2*C^2*H*V*a^2*b*c*y2(t)_0^2 + C^2*H*a*c*y1(t)_0*y2(t)_0^2
+ 3*C^2*V*a^2*b*c*y2(t)_0^2*y3(t)_0 + C*H^2*V*a^2*b*c*y2(t)_0
- C*H^2*a*c*y1(t)_0*y2(t)_0 + C*H^2*gamma*y1(t)_0*y2(t)_0 + C*H^2*mu*y1(t)_0*y2(t)_0
+ C*H^2*y2(t)_0*y1(t)_1 - 4*C*H*V*a^2*b*c*y2(t)_0*y3(t)_0
+ 2*C*H*a*c*y1(t)_0*y2(t)_0*y3(t)_0 + 3*C*V*a^2*b*c*y2(t)_0*y3(t)_0^2
- H^3*mu*y1(t)_0 - H^3*y1(t)_1
+ H^2*V*a^2*b*c*y3(t)_0 - H^2*a*c*y1(t)_0*y3(t)_0 + H^2*gamma*y1(t)_0*y3(t)_0
+ H^2*mu*y1(t)_0*y3(t)_0 - H^2*y1(t)_0^2 + H^2*y3(t)_0*y1(t)_1
- 2*H*V*a^2*b*c*y3(t)_0^2 + H*a*c*y1(t)_0*y3(t)_0^2 + V*a^2*b*c*y3(t)_0^3
```

which can be rewritten as

$$\begin{aligned}
&C^3Va^2bcy_2^3 - 2C^2HVa^2bcy_2^2 + C^2Hacy_1y_2^2 + 3C^2Va^2bcy_2^2y_3 \\
&+ CH^2Va^2bcy_2 - CH^2acy_1y_2 + CH^2\gamma y_1y_2 + CH^2\mu y_1y_2 + CH^2y_2y_1' \\
&- 4CHVa^2bcy_2y_3 + 2CHacy_1y_2y_3 + 3CVa^2bcy_2y_3^2 - H^3\mu y_1 - H^3y_1' \\
&+ H^2Va^2bcy_3 - H^2acy_1y_3 + H^2\gamma y_1y_3 + H^2\mu y_1y_3 - H^2y_1^2 + H^2y_3y_1' \\
&- 2HVa^2bcy_3^2 + Hacy_1y_3^2 + Va^2bcy_3^3 = 0.
\end{aligned} \tag{S7.6}$$

Dividing (S7.6) by $H^2$ gives:

$$\begin{aligned}
&\frac{C^3Va^2bcy_2^3}{H^2} - \frac{2C^2Va^2bcy_2^2}{H} + \frac{C^2acy_1y_2^2}{H} + \frac{3C^2Va^2bcy_2^2y_3}{H^2} \\
&+ CVa^2bcy_2 - Cacy_1y_2 + C\gamma y_1y_2 + C\mu y_1y_2 + Cy_2y_1' \\
&- \frac{4CVa^2bcy_2y_3}{H} + \frac{2Cacy_1y_2y_3}{H} + \frac{3CVa^2bcy_2y_3^2}{H^2} - H\mu y_1 - Hy_1' \\
&+ Va^2bcy_3 - acy_1y_3 + \gamma y_1y_3 + \mu y_1y_3 - y_1^2 + y_3y_1' \\
&- \frac{2Va^2bcy_3^2}{H} + \frac{acy_1y_3^2}{H} + \frac{Va^2bcy_3^3}{H^2} = 0,
\end{aligned} \tag{S7.7}$$

from which it follows that (for example)

$$C = C^\star, \quad C\gamma = C^\star\gamma^\star, \quad C\mu = C^\star\mu^\star, \quad H\mu = H^\star\mu^\star, \quad Cac = C^\star a^\star c^\star, \quad \text{and} \quad Va^2bc = V^\star(a^2)^\star b^\star c^\star,$$

which implies that $C$ alongside $\gamma, \mu, H, ac$, and $Vab$ are globally identifiable. The fact that $V$, $a$, $b$, and $c$ are nonidentifiable can be established in the same way as in Section S4(i) by using the transformation $V \to \lambda V,\ a \to \lambda a,\ b \to \lambda b$.

#### (ii) Identifiability of state variables

With $C$ and $\gamma$ proven to be globally identifiable, it follows from (S7.5) that $I_h(t)$ is also globally identifiable. Since $I_h$ is globally identifiable, global identifiability of $X$ follows from (S7.3) as $a_2$ and $H$ are globally identifiable while $y$ is the observable.

### S8 Proof of Theorem 3.8

*Proof.* For convenience, we rewrite the behavior model (3.9) with saturation exponent $k = 1$:

$$\begin{cases} S' = -\dfrac{\beta S I}{1+\alpha I}, \\ I' = \dfrac{\beta S I}{1+\alpha I} - \gamma I, \\ R' = \gamma I, \end{cases} \tag{S8.1}$$

#### (i) Identifiability of parameters

Julia gives the following input-output equation:

```
-alpha*gamma*y(t)_0^2*y(t)_1 - alpha*y(t)_0^2*y(t)_2 - beta*gamma*y(t)_0^3
- beta*y(t)_0^2*y(t)_1 - y(t)_0*y(t)_2 + y(t)_1^2
```

which can be rewritten as

$$-\alpha\gamma y^2 y' - \alpha y^2 y'' - \beta\gamma y^3 - \beta y^2 y' - y y'' + (y')^2 = 0 \tag{S8.2}$$

The following input-output equation is obtained from DAISY:

```
df(y1,t,2)*y1**3*alpha**2 + 2*df(y1,t,2)*y1**2*alpha + df(y1,t,2)*y1
- df(y1,t)**2*y1*alpha - df(y1,t)**2 + df(y1,t)*y1**3*alpha*(alpha*gamma + beta)
+ df(y1,t)*y1**2*(alpha*gamma + beta) + y1**4*alpha*beta*
gamma + y1**3*beta*gamma$
```

which can be rewritten as

$$y''y^3\alpha^2 + 2y''y^2\alpha + y''y - (y')^2 y\alpha - (y')^2 + y'y^3\alpha(\alpha\gamma+\beta) + y'y^2(\alpha\gamma+\beta) + y^4\alpha\beta\gamma + y^3\beta\gamma = 0 \tag{S8.3}$$

The equations (S8.2) and (S8.3) are not identical. However, one can verify that the latter can be factored as

$$-(1+\alpha y)(-\alpha\gamma y^2 y' - \alpha y^2 y'' - \beta\gamma y^3 - \beta y^2 y' - y y'' + (y')^2) = 0,$$

where the second factor is precisely (S8.2). Since $1 + \alpha y$ does not vanish in every solution of (S8.1), the input-output relation obtained by DAISY in fact implies the one obtained by StructuralIdentifiability.jl thus confirming the correctness of (S8.2). Furthermore, (S8.2) cannot be factored further, so it is minimal. Hence, it is (S8.2) which should be used for assessing identifiability [8, Section 4].

Combining like terms in (S8.2) and dividing by $-\alpha$ yields the following monic polynomial:

$$y^2 y'' + \frac{\alpha\gamma+\beta}{\alpha} y^2 y' + \frac{\beta\gamma}{\alpha} y^3 + \frac{1}{\alpha} y y'' - \frac{1}{\alpha}(y')^2 = 0 \tag{S8.4}$$

The coefficients of this normalized equation are identifiable, so we obtain global identifiability of $\alpha$, $\beta\gamma = \left(\frac{\beta\gamma}{\alpha}\right)\alpha$, and $\alpha\gamma + \beta = \left(\frac{\alpha\gamma+\beta}{\alpha}\right)\alpha$ as claimed in the theorem. In order to show that $\beta$ and $\gamma$ are locally identifiable, we observe that $\alpha\gamma$ and $\beta$ are roots of an equation $X^2 - (\alpha\gamma+\beta)X + \alpha\gamma\beta = 0$ with identifiable coefficients, so they are locally identifiable. Local identifiability of $\alpha\gamma$ and global identifiability of $\alpha$ now imply local identifiability of $\gamma = \frac{\alpha\gamma}{\alpha}$.

Finally, in order to show that $\beta$ and $\gamma$ are not locally identifiable, we provide the following parameter transformation which keeps invariant the coefficients of the input-output equations:

$$\beta \to \alpha\gamma, \quad \gamma \to \frac{\beta}{\alpha}. \tag{S8.5}$$

This transformation allows to construct, for the values $(\alpha, \beta, \gamma)$ another vector with different $\beta$ and $\gamma$ but the same input-output equation. This, $\beta$ and $\gamma$ are not globally identifiable.

#### (ii) Identifiability of state variables

We reparameterize model (S8.1) through the following parameter and state variable and transformations:

$$(a_1, a_2) = (\beta\gamma, \alpha\gamma + \beta) \quad \text{and} \quad (X_1, X_2) = (\beta S + \beta I - \gamma, I),$$

to obtained the following reparameterized model:

$$\begin{cases} X_1' = -a_1 X_2, \\ X_2' = \dfrac{X_1 X_2 - a_2 X_2^2}{1 + \alpha X_2} \end{cases} \tag{S8.6}$$

Notice that $a_1$ and $a_2$ are globally identifiable. Given $X_2 = I$ is the observation it follows that $X_2'$ is also known. Moreover, since $\alpha, a_1$, and $a_2$ are identifiable, it follows from the second equation in (S8.6) that $X_2$ is globally identifiable.

Hence, $I$ and $\beta(S+I)-\gamma$ are globally identifiable. Note that $S$ can be written in terms of locally identifiable quantities

$$S = \frac{(\beta(S+I)-\gamma)+\gamma}{\beta} - I,$$

so it is locally identifiable. In order to show that $S$ is not globally identifiable, we extend the transformation (S8.5) to an output-preserving transformation involving states and parameters:

$$\beta \to \alpha\gamma, \quad \gamma \to \frac{\beta}{\alpha}, \quad S \to \frac{\beta(S+I) - \gamma + \frac{\beta}{\alpha}}{\alpha\gamma} - I.$$

Direct substitution shows that this transformation does not change the output but changes the initial condition for $S(t)$ thus showing that it is not globally identifiable.

#### (iii) Identifiability with one additional complementary data point

Since $\alpha$ and $\beta(S+I)-\gamma$ are globally identifiable, so is $\alpha(\beta(S+I)-\gamma)$. Moreover, since $\alpha\gamma+\beta$ is also globally identifiable, so is $\alpha(\beta(S+I)-\gamma)+(\alpha\gamma+\beta)$, which can be rewritten as:

$$\alpha(\beta(S+I)-\gamma)+(\alpha\gamma+\beta) = \beta(\alpha(S+I)+1),$$

which implies $\beta(\alpha(S+I)+1)$ must be globally identifiable. Since $I(t)$ is the observable and $\alpha$ is globally identifiable, it follows that having a single data point in $S(t)$ yields identifiability of $\beta$. Since $\beta\gamma$ is globally identifiable, identifiability of $\beta$ yields identifiability of $\gamma$, thus making all parameters globally identifiable. Moreover, with $\beta$ and $\gamma$ identifiable, the fact that $\beta(S+I)-\gamma$ is identifiable, yields in global identifiability of $S$. Finally, with $S$ identifiable, $R = N - S - I$ is also globally identifiable. Consequently, the entire model is globally identifiable. □

#### S9 Idea Behind Conjecure 2

For convenience, we rewrite behavior model (3.9):

$$\begin{cases} S' = -\dfrac{\beta SI}{(1+\alpha I)^k}, \\ I' = \dfrac{\beta SI}{(1+\alpha I)^k} - \gamma I, \\ R' = \gamma I, \end{cases} \tag{S9.1}$$

Since $R'$ equation is decoupled, we rewrite model (S9.1) as

$$\begin{cases} S' = -\dfrac{\beta SI}{(1+\alpha I)^k}, \\ I' = \dfrac{\beta SI}{(1+\alpha I)^k} - \gamma I, \end{cases} \tag{S9.2}$$

Let $y = I$ be the observation.

The intuition behind Conjecture 2 is given below for $k = 2, 3, 4, 5, 10, 50$ using `StructuralIdentifiability.jl`. For $k = 1$, model (S2.1) reduces to the SIR model (3.1).

***k* = 2**

```
ode = @ODEmodel(
    S'(t) = - beta*S(t)*I(t)//(1 + alpha*I(t))^2,
    I'(t) = beta*S(t)*I(t)//(1 + alpha*I(t))^2 - gamma*I(t),
    y(t) = I(t)
)
```

`assess_identifiability(ode)` gives:

```
S(t)  => :globally
I(t)  => :globally
alpha => :globally
beta  => :globally
gamma => :globally
```

Global identifiability of all parameters and state variables $S(t)$ and $I(t)$ is also obtained for $k = 3, 4, 5, 10$ and 50. Since $N(0) = S(0) + I(0) + R(0)$ is assumed to be known and the population is closed, $N(t) = S(t) + R(t) + S(t)$ with $N(t) = N(0)$. Thus, global identifiability of $S(t)$ and $I(t)$, together with $N(t)$ being known, also implies that $R(t)$ is known. Hence, the model appears to be globally identifiable for $k \geq 2$.

While a proof of global identifiability for arbitrary $k > 1$ remains elusive, we propose an alternative formulation of the model that yields global identifiability for *large enough k*.

**Remark 1** (Global identifiability for large enough $k$)**.** We introduce a new variable $A := \frac{(1+\alpha I)^k}{\beta}$. Then, in terms of $A$, we have

$$\begin{cases} S' = -\dfrac{SI}{A}, \\ I' = \dfrac{SI}{A} - \gamma I, \\ A' = k\alpha I' \dfrac{(1+\alpha I)^k}{\beta} = \dfrac{k\alpha I' A}{1+\alpha I} = \dfrac{\alpha k I(S - \gamma A)}{1+\alpha I}. \end{cases} \tag{S9.3}$$

Direct computation with `StructuralIdentifiability.jl` shows that all the parameters and states in this model (S9.3) are globally identifiable. This implies that the values of parameters and initial conditions can be expressed as rational functions in $y(0), y'(0), y''(0), \ldots$ by [6, Proposition 3.4]. The denominators of these rational functions may vanish only at finitely many values of $k$, so , for large enough value of $k$ (the one which does not annihilate any of the denominators of these rational functions), the specialization of the model at this $k$ is globally identifiable as well. For every such $k$, since we can recover $\beta$ as $\frac{(1+\alpha I)^k}{A}$, $\beta$ is also globally identifiable, so the original model is globally identifiable as well.

**Remark 2** (What is special about $k = 1$?)**.** As we have already highlighted, the model (3.9) is a nice illustration of the fact that identifiability patterns may be unexpected and non-obvious. In hindsight, one general idea behind the observed change between $k = 1$ and $k > 1$ is that linear (or close to linear) models tend to exhibit nontrivial nonidentifiability behavior in a more systematic fashion (see, e.g. [1, Appendices] and [5]). This may be explained by the fact that the input-output equations for such models are linear as well and, thus, do not contain too many coefficients. For example, the number of nonconstant coefficients in the input-output equation of a linear single-output model of dimension $n$ is at most $n$. Thus, if such a model contains more than $n$ parameters, it cannot be globally identifiable simply for dimension reasons.

For reference, the input-output equation, obtained using `StructuralIdentifiability.jl`, for $k = 2, 3, 4$ is given below. $k = 2$

```
-2*alpha^2*gamma*y(t)_0^3*y(t)_1 - alpha^2*y(t)_0^3*y(t)_2
- alpha^2*y(t)_0^2*y(t)_1^2 - 2*alpha*gamma*y(t)_0^2*y(t)_1 - 2*alpha*y(t)_0^2*y(t)_2
- beta*gamma*y(t)_0^3 - beta*y(t)_0^2*y(t)_1 - y(t)_0*y(t)_2 + y(t)_1^2
```

$k = 3$

```
-3*alpha^3*gamma*y(t)_0^4*y(t)_1 - alpha^3*y(t)_0^4*y(t)_2
- 2*alpha^3*y(t)_0^3*y(t)_1^2 - 6*alpha^2*gamma*y(t)_0^3*y(t)_1
- 3*alpha^2*y(t)_0^3*y(t)_2 - 3*alpha^2*y(t)_0^2*y(t)_1^2
- 3*alpha*gamma*y(t)_0^2*y(t)_1 - 3*alpha*y(t)_0^2*y(t)_2 - beta*gamma*y(t)_0^3
- beta*y(t)_0^2*y(t)_1 - y(t)_0*y(t)_2 + y(t)_1^2
```

$k = 4$

```
-4*alpha^4*gamma*y(t)_0^5*y(t)_1 - alpha^4*y(t)_0^5*y(t)_2
- 3*alpha^4*y(t)_0^4*y(t)_1^2 - 12*alpha^3*gamma*y(t)_0^4*y(t)_1
- 4*alpha^3*y(t)_0^4*y(t)_2 - 8*alpha^3*y(t)_0^3*y(t)_1^2
- 12*alpha^2*gamma*y(t)_0^3*y(t)_1 - 6*alpha^2*y(t)_0^3*y(t)_2
- 6*alpha^2*y(t)_0^2*y(t)_1^2 - 4*alpha*gamma*y(t)_0^2*y(t)_1
- 4*alpha*y(t)_0^2*y(t)_2 - beta*gamma*y(t)_0^3 - beta*y(t)_0^2*y(t)_1
- y(t)_0*y(t)_2 + y(t)_1^2
```

which can be respectively written as:

$$-2\alpha^2\gamma y^3 y' - \alpha^2 y^3 y'' - \alpha^2 y^2 (y')^2 - 2\alpha\gamma y^2 y' - 2\alpha y^2 y'' - \beta\gamma y^3 - \beta y^2 y' - y y'' + (y')^2 = 0,$$

$$-3\alpha^3\gamma y^4 y' - \alpha^3 y^4 y'' - 2\alpha^3 y^3 (y')^2 - 6\alpha^2\gamma y^3 y' - 3\alpha^2 y^3 y'' - 3\alpha^2 y^2 (y')^2 - 3\alpha\gamma y^2 y' - 3\alpha y^2 y'' - \beta\gamma y^3 - \beta y^2 y' - y y'' + (y')^2 = 0,$$

$$\begin{aligned} -4\alpha^4\gamma y^5 y' - \alpha^4 y^5 y'' - 3\alpha^4 y^4 (y')^2 - 12\alpha^3\gamma y^4 y' - 4\alpha^3 y^4 y'' - 8\alpha^3 y^3 (y')^2 - 12\alpha^2\gamma y^3 y' \\ - 6\alpha^2 y^3 y'' - 6\alpha^2 y^2 (y')^2 - 4\alpha\gamma y^2 y' - 4\alpha y^2 y'' - \beta\gamma y^3 - \beta y^2 y' - y y'' + (y')^2 = 0. \end{aligned}$$
